# Risk of major adverse cardiovascular events with dolutegravir versus efavirenz-based antiretroviral therapy: emulated target trials using routine, de-identified data from South Africa

**DOI:** 10.1101/2025.03.07.25323562

**Authors:** Jienchi Dorward, Xolani Masombuka, Lara Lewis, Claudia Pastellides, Johan van der Molen, Kwabena Asare, Kwena Tlhaku, Jennifer Anne Brown, Christian Bottomley, Dave Jacobs, Shirley Collie, Nigel Garrett

## Abstract

**Background:** Integrase inhibitors, including dolutegravir, may increase risk of major adverse cardiovascular events (MACEs). However, limited data exists from low- and middle-income countries, where tenofovir disoproxil fumarate, lamivudine and dolutegravir (TLD) has largely replaced tenofovir disoproxil fumarate, emtricitabine and efavirenz (TEE).

**Methods:** We used de-identified data from a South African managed-healthcare organisation from people living with HIV (PLHIV) without cardiovascular disease, who either initiated TEE or TLD between April 2020-Dec 2023 (initiation cohort) or were receiving TEE in April 2020 and eligible for TLD (transition cohort). In the initiation cohort, we emulated a target trial using pooled logistic regression models with inverse probability of treatment weights and bootstrapped confidence intervals to compare standardised 3-year MACE risk between TLD versus TEE. In the transition cohort, we used similar methods in 44 emulated monthly sequential trials, comparing MACE risk in people transitioned to TLD with those remaining on TEE.

**Findings:** In the initiation cohort, 7310 PLHIV initiated TLD (n=3711) or TEE (n=3599). Median follow-up was 21 months (IQR 10-33), with 18 MACEs with TLD (3-year risk 0.78%, 95%CI 0.37-1.32) and 28 with TEE (3-year risk 0.96%, 0.60-1.40; RR 0.81, 0.35-1.59; RD −0.18, −0.82-0.50). In the transition cohort, 22338 people contributed to 2837 person-trials with TLD and 706615 with TEE. Median follow-up was 25 months (14-36), with 19 MACEs with TLD (3-year risk 1.09%, 0.48-1.99) and 5420 with TEE (3-year risk 1.21%, 1.05-1.41; RR 0.90, 0.41-1.64; RD −0.12, −0.75-0.75).

**Interpretation:** Among PLHIV in South Africa we found no increased MACE with TLD.

**Funding:** Gates Foundation; National Institute of Health and Social Care Research

**RESEARCH IN CONTEXT:** *Evidence before this study:* We searched PubMed with no language restrictions on March 6^th^, 2025, with the terms “(dolutegravir) AND (cardiovascular disease OR coronary heart disease OR cerebrovascular disease OR stroke)” and identified additional studies using hand searches of reference lists and citing papers. We found no randomised trials which were adequately powered to directly assess the risk of major adverse cardiovascular events (MACEs) between dolutegravir (or integrase strand transferase inhibitors [INSTIs]) and efavirenz (or non-nucleoside reverse transcriptase inhibitors). We identified one systematic review from 2018 of eight trials, predominantly from high-income settings, which found 15/2202 (0.7%) serious adverse cardiovascular events with dolutegravir versus 8/2215 (0.4%) with other antiretrovirals (relative risk 1.69, 95% CI 0.71 to 4.03). We identified five observational studies which assessed risk of cardiovascular events with INSTIs versus non-INSTI antiretroviral therapy (ART). A study using medical insurance claims data from the United States between 2008 and 2015 found initiating an INSTI was associated with fewer cardiovascular events compared to non-INSTI initiation, while a later study using the same dataset from 2013 to 2021 found no difference in MACE between INSTI versus non-INSTI initiation, although INSTI use was associated with increased myocardial infarction. An observational study using 17 European and Australian cohorts found an association between cumulative INSTI exposure up to 24 months and increased risk of cardiovascular events, although the study design has been questioned. Two studies used observational data to emulate target trials comparing risk of cardiovascular events among people using INSTI versus non-INSTI ART. In a Swiss cohort, people initiating INSTIs were not found to be at increased risk of cardiovascular events, while in a larger study using data from European and North American cohorts, 4-year cardiovascular risk was similar between INSTI and non-INSTI users in both ART naïve and ART experienced individuals.

*Added value of this study:* Our study is the first to evaluate risks of MACEs with tenofovir disoproxil fumarate, lamivudine and dolutegravir (TLD), the most widely used INSTI-based regimen in low- and middle-income countries (LMICs), where the majority of people living with HIV (PLHIV) live. This is important as this regimen has been recommended by the World Health Organisation (WHO) for first-line ART since 2018, replacing the previously recommended regimen of tenofovir disoproxil fumarate, emtricitabine and efavirenz (TEE). Using robust emulated target trial methods, we found no evidence of increased risk of MACEs with TLD versus TEE in both people initiating ART, or people already ART-experienced, in a large South African cohort. These findings are relevant for the over 20 million people estimated to be taking TLD in LMICs, where risk factors for cardiovascular disease may differ from high-income settings.

*Implications of all the available evidence:* We found no large increased risk of MACEs in the short-to medium term with TLD, which is supported by the majority of evidence investigating risks with INSTIs from high-income settings. These findings support the ongoing use of dolutegravir-based ART as part of the WHO public health approach in LMICs, although studies with greater follow-up time are required.

## INTRODUCTION

The integrase strand transferase inhibitor (INSTI) dolutegravir is recommended for first- and second-line antiretroviral therapy (ART) in over 118 low- and middle-income countries (LMICs), and is used by over 20 million people living with HIV (PLHIV).^1,2^ Dolutegravir has better efficacy, fewer side effects and a higher genetic barrier to resistance, compared to the previously recommended efavirenz.^3^ However, there are concerns regarding a potential association between INSTI use and major adverse cardiovascular events (MACEs), with an observational study in European and Australian cohorts finding increased MACE risk in the first 24 months of INSTI use.^4^ Furthermore, several African clinical trials found that dolutegravir was associated with greater weight gain than efavirenz, particularly among women,^5,6^ although whether this translates into increased MACE risk remains unclear. Findings from observational studies, which have tended to focus on INSTIs as a group rather than dolutegravir alone, have been mixed,^4,7–10^ and conducted predominantly in European or North American populations. In African populations, which contain the largest number of people taking dolutegravir, studies have not been sufficiently powered to evaluate MACEs.^11,12^

We aimed to assess whether dolutegravir increases MACE risk compared to the previously recommended efavirenz among PLHIV in South Africa.

## METHODS

We used observational data to emulate target trials, a methodology that aims to reduce bias when using observational data for causal inference.^13,14^ Following reporting recommendations,^15^ we specify key components of the hypothetical target trials that we aimed to emulate, before describing the observational data and emulation methods.

### Target trial specifications

We emulated target trials in two cohorts, people initiating ART (initiation cohort), and people already receiving first-line ART (transition cohort) (Table 1). For the initiation target trial, eligible participants would be PLHIV aged _≥_18 years, without known cardiovascular disease (CVD), and newly initiating ART, and would be randomised at baseline to initiate open-label tenofovir disoproxil fumarate, lamivudine and dolutegravir (TLD) or tenofovir disoproxil fumarate, emtricitabine and efavirenz (TEE) (Table 1). For the transition target trial, eligible participants would be PLHIV aged _≥_18 years already receiving TEE, without current viraemia >1000 copies/mL, without known CVD, and eligible for transition to first-line TLD. People would be randomised at baseline to either continue TEE, or be transitioned to TLD. In both trials, the primary outcome, MACE (cardiovascular death or hospitalization), would be assessed over 36 months, with censoring at ART gap >6 months, death, withdrawal, MACE, or study end. The primary analysis would be an intention-to-treat analysis, with a secondary per-protocol analysis. Because the effect of dolutegravir on weight gain is greater among women,^5,6^ and the excess risk of MACE due to HIV may be greater among women,^16^ we planned a sensitivity analysis to examine sub-group effects by gender on the risk of MACE with TLD. The 3-year standardised cumulative risk of MACE in each arm would be estimated using hazards estimated using pooled logistic regression models and compared using risk ratios and risk differences, with 95% confidence intervals calculated using 500 bootstrap samples. In the per-protocol analysis, participants would additionally be censored if they change ART, and the pooled logistic regression model would be weighted for the inverse probability of any censoring, estimated using baseline and time varying co-variates.

**Table 1:** Specification of the target trials.

|  | Initiation cohort analysis | Transition cohort analysis |
| --- | --- | --- |
| Eligibility criteria | Person living with HIV aged $\geq 18$ years old, without previous known cardiovascular disease, and newly initiating ART | Person living with HIV aged $\geq 18$ years old, already receiving TEE, without current viraemia ($>1000$ copies/mL), without previous known cardiovascular disease, and eligible for transition to first-line TLD |
| Treatment strategies | Initiated on TLD or TEE | Transition immediately to TLD or continue TEE |
| Treatment assignment | Randomised, with clinician and participant aware of allocation |  |
| Outcome | Major adverse cardiovascular events (MACE) consisting of cardiovascular death or hospitalisation |  |
| Follow-up | Followed up until the earliest of treatment interruption, death, withdrawal, MACE, or 36 months |  |
| Causal contrasts of interest | Primary analysis would be an intention to treat analysis, with a secondary per-protocol analysis |  |
| Analysis plan | Pooled logistic regression model to estimate time-varying hazards, which are used to calculate the standardised 3 year risk of MACE under TLD versus TEE, compared with risk ratios and risk differences | Pooled logistic regression model, to estimate time-varying hazards, which are used to calculate the standardised 3 year risk of MACE under TLD versus TEE, compared with risk ratios and risk differences. |

### Observational data source and data management

We used de-identified, routinely collected data from a South African managed-healthcare organisation (Discovery Health, Johannesburg, South Africa), that collects and securely processes healthcare data on members for the purposes of administering medical aid schemes and funding healthcare service provision. In this scheme, ART is provided through private general practitioners or infectious diseases specialists, normally following Southern African HIV Clinician Society or South African National Department of Health guidelines.^17,18^ Dolutegravir started to be widely used for first-line ART from early 2020, when it was introduced into the public sector in a fixed dose combination pill of TLD, replacing a fixed dose combination pill of TEE. Initially, TLD use was restricted in women of child-bearing potential due to safety concerns, but in July 2021, this recommendation was lifted, and TLD became the preferred first-line regimen.^19^ Viral load testing was recommended 6-monthly or annually, and transition to first-line dolutegravir was only recommended if people had a suppressed viral load of <50 copies/mL in the previous six months, or consecutive viral loads between 50-999 copies/mL.^17^ After July 2022, these viral suppression criterion were removed.^20^

For all members, the managed-healthcare organisation collects and processes data on self-reported medical conditions at enrolment, new diagnoses during follow-up, claims for medication prescriptions (including ART), hospitalisation diagnostic codes, laboratory investigations and results, and cause of death. Data on confirmed chronic conditions, verified through physician documentation and valid claims containing appropriate diagnosis codes, was collected, cleaned, cross-checked between different databases, anonymised, and securely processed by the managed-healthcare organisation prior to extraction for analysis.

### Participants

For the initiation cohort, we included PLHIV aged _≥_18 years, without known CVD, and newly initiating TLD or TEE first-line ART within the managed-healthcare cohort, between April 1, 2020 and December 31, 2023. This allowed at least six months of follow-up and three months of data capture ‘run-off’ before the data cut on September 30, 2024. We excluded people with known previous ART exposure, <6-months of insurance scheme membership (as it was not possible to know if they joined the scheme while already receiving ART) or suppressed viral load <1000 copies/mL at initiation, which may suggest current or recent ART exposure. For the transition cohort, we included PLHIV aged _≥_18 years, without known CVD, and already receiving TEE in the managed-healthcare cohort in April 2020, and followed them until June 30, 2024, again to allow three-months of data capture ‘run-off’ before the data cut.

### Variables

#### Outcomes

We used cause of death and hospital admission codes to define the primary endpoint of MACE as a composite of death related to acute myocardial infarction or stroke, or hospital admission for acute myocardial infarction, unstable angina, stroke (ischemic, haemorrhagic or undetermined), transient ischemic attack, peripheral arterial ischemia and coronary, carotid or peripheral artery revascularisation (e.g. angioplasty, stenting, coronary bypass surgery, carotid endarterectomy). People who withdrew from the medical scheme were defined as lost to follow up on the date of withdrawal.

#### Exposure variable

We used ART claims data to determine ART exposure at baseline and during follow-up. We used the first TLD or TEE claim to determine the date of initiation (initiation cohort), and the most recent claim in April 2020 to determine baseline ART exposure in the transition cohort. We censored anyone with a gap in ART claims >6-months due to uncertainty in ART exposure and continued scheme activity regarding claims for hospitalisations.

#### Covariates

At baseline and throughout follow-up we used laboratory data to determine CD4 T-cell counts and viral loads, and chronic illness benefit application forms and disease specific claims to determine CVD, hypercholesterolaemia, hypertension, diabetes mellitus, pregnancy and tuberculosis episodes. We determined statin use using prescription claims, and used scheme benefit level as a proxy for socioeconomic status. Where data was missing, we included a category for missing data.

### Statistical analysis

In the initiation cohort intention-to-treat analysis, we emulated randomisation between TLD or TEE using stabilized inverse probability of treatment weights (IPTWs), calculated using propensity scores from a logistic regression model with treatment assignment as the outcome and potential confounders at baseline as covariates (age, gender, province, scheme benefit level, initiation period [in quarterly intervals], CD4 count, viral load, known TB, known pregnancy, known diabetes, known hypertension, known hypercholesterolaemia, statin use). We then estimated the hazards of MACE by fitting an IPTW pooled logistic regression model with a time-varying intercept, a treatment assignment variable and a treatment-time interaction. We used the model to predict monthly outcomes under the scenarios of all participants receiving TEE, and all participants receiving TLD, and calculated the standardised 3-year risk of MACE with TEE and TLD, the risk ratio and risk difference, with 95% confidence intervals estimated from 500 bootstrap samples. For the per-protocol analysis, we used a similar approach, but with stabilized inverse probability of censoring weights (IPCWs) in the final outcome model, calculated using a pooled logistic regression model for the monthly risk of censoring, with baseline (ART regimen and the same variables as the IPTW model) and time-updated variables (follow-up time in months, viral load, CD4, statin use, pregnancy status, and incident tuberculosis, diabetes mellitus, hypercholesterolaemia and hypertension) as covariates.

For the transition cohort intention-to-treat analysis, we emulated 44 sequential target trials,^21^ each using a different baseline month from May 2020 to December 2023. We emulated randomisation, between remaining on TEE versus transitioning in the baseline month to TLD, using IPTW, calculated using covariate values from baseline of the respective trial (age, gender, province, scheme benefit level, baseline CD4 count, baseline viral load, TB status, pregnancy status, known diabetes, known hypertension, known hypercholesterolaemia, statin use). As per the target trial eligibility criteria, people who were transitioned to TLD were excluded from subsequent trials, but people who remained on TEE could have been included in subsequent trials, meaning individuals could appear in multiple trials. We then used similar methods to the initiation cohort analysis to estimate the standardised 3-year risk of MACE under TLD and TEE, and the risk difference and risk ratio, with 500 bootstrap samples to estimate 95% confidence intervals. For the per protocol analysis individuals within each trial were additionally censored upon ART regimen changes, and the outcome model again included IPCWs.

In both the initiation and transition analyses we conducted sensitivity analyses including body-mass index (with a category for missing) in the IPTW (and IPCW) models, and further sensitivity analyses with an interaction term between treatment assignment and gender in the outcome model.

We analysed data using R 4.4.0 (R Foundation for Statistical Computing, Vienna, Austria), with code available in the supplementary appendix.

### Ethical approval

This work was approved by the University of Kwazulu-Natal Biomedical Research Ethics Committee (BREC/00005858/2023), with a waiver for informed consent for analysis of de-identified, routinely collected data.

## RESULTS

### Initiation cohort

In the initiation cohort, between April 1^st^, 2020 and December 31^st^, 2023, 7310 people initiated TLD (n=3711) and TEE (n=3599). Median (IQR) age was 38 (32-44) years, 57.0% were female, and 14.2% had a recorded CVD risk factor (Table 2). Baseline body-mass index (BMI) was available for 1993 (27.3%) of participants; of these 1394 (70.0%) were overweight, obese or severely obese. The TLD group had fewer women (54.4% versus 59.8%), pregnant people (5.3% versus 10.8%) and people with missing baseline BMI (66.3% versus 79.4%) versus TEE. However, among those with BMI recorded, distributions were similar. There was a higher proportion of people who were initiated later in the study period (e.g. Oct-Dec 2023 8.8% versus 2.3%) in the TLD versus TEE groups. After IPTW, baseline covariates were well balanced between the two groups (Table 2).

**Table 2:** Baseline characteristics of initiation cohort, and emulated target trial pseudo-population after inverse probability of treatment weighting.

| Variable | Level | Initiation cohort |  |  | Target trial pseudo-population |  |  |
| --- | --- | --- | --- | --- | --- | --- | --- |
|  |  | TEE (n = 3599, 49.2%) | TLD (n = 3711, 50.8%) | Total (n = 7310) | TEE (n=3622) | TLD (n=3689) | Standardised mean difference |
| Age, years | Mean (SD) | 38.7 (8.9) | 39.0 (8.9) | 38.8 (8.9) | 38.90 (8.86) | 39.01 (9.02) | 0.012 |
| Gender | Female (%) | 2151 (59.8) | 2017 (54.4) | 4168 (57.0) | 2047.0 (56.5) | 2078.0 (56.3) | 0.004 |
| Province | GAUTENG | 1502 (41.7) | 1665 (44.9) | 3167 (43.3) | 1555.6 (42.9) | 1590.0 (43.1) | 0.012 |
|  | EASTERN CAPE | 223 (6.2) | 232 (6.3) | 455 (6.2) | 220.0 ( 6.1) | 221.3 ( 6.0) |  |
|  | KWA ZULU NATAL | 793 (22.0) | 697 (18.8) | 1490 (20.4) | 743.6 (20.5) | 750.7 (20.3) |  |
|  | MPUMALANGA | 277 (7.7) | 284 (7.7) | 561 (7.7) | 288.9 ( 8.0) | 298.1 ( 8.1) |  |
|  | OTHER | 558 (15.5) | 537 (14.5) | 1095 (15.0) | 536.6 (14.8) | 555.0 (15.0) |  |
|  | WESTERN CAPE | 246 (6.8) | 296 (8.0) | 542 (7.4) | 277.2 ( 7.7) | 274.1 ( 7.4) |  |
| Initiation time period | Apr - Jun 2020 | 338 (9.4) | 118 (3.2) | 456 (6.2) | 225.2 ( 6.2) | 237.2 ( 6.4) | 0.017 |
|  | Jul - Sep 2020 | 244 (6.8) | 104 (2.8) | 348 (4.8) | 169.6 ( 4.7) | 165.7 ( 4.5) |  |
|  | Oct - Dec 2020 | 277 (7.7) | 115 (3.1) | 392 (5.4) | 191.8 ( 5.3) | 190.9 ( 5.2) |  |
|  | Jan - Mar 2021 | 653 (18.1) | 267 (7.2) | 920 (12.6) | 452.0 (12.5) | 462.3 (12.5) |  |
|  | Apr - Jun 2021 | 286 (7.9) | 190 (5.1) | 476 (6.5) | 233.3 ( 6.4) | 237.5 ( 6.4) |  |
|  | Jul - Sep 2021 | 266 (7.4) | 189 (5.1) | 455 (6.2) | 223.2 ( 6.2) | 226.8 ( 6.1) |  |
|  | Oct - Dec 2021 | 222 (6.2) | 161 (4.3) | 383 (5.2) | 187.8 ( 5.2) | 194.3 ( 5.3) |  |
|  | Jan - Mar 2022 | 256 (7.1) | 252 (6.8) | 508 (6.9) | 250.4 ( 6.9) | 256.6 ( 7.0) |  |
|  | Apr - Jun 2022 | 186 (5.2) | 239 (6.4) | 425 (5.8) | 207.2 ( 5.7) | 214.0 ( 5.8) |  |
|  | Jul - Sep 2022 | 171 (4.8) | 312 (8.4) | 483 (6.6) | 240.5 ( 6.6) | 246.3 ( 6.7) |  |
|  |  | TEE (n = 3599, 49.2%) | TLD (n = 3711, 50.8%) | Total (n = 7310) | TEE (n=3622) | TLD (n=3689) | Standardised mean difference |
|  | Oct - Dec 2022 | 184 (5.1) | 348 (9.4) | 532 (7.3) | 270.1 ( 7.5) | 273.6 ( 7.4) |  |
|  | Jan - Mar 2023 | 182 (5.1) | 379 (10.2) | 561 (7.7) | 277.8 ( 7.7) | 284.5 ( 7.7) |  |
|  | Apr - Jun 2023 | 133 (3.7) | 327 (8.8) | 460 (6.3) | 227.2 ( 6.3) | 234.3 ( 6.4) |  |
|  | Jul - Sep 2023 | 119 (3.3) | 384 (10.3) | 503 (6.9) | 259.6 ( 7.2) | 257.5 ( 7.0) |  |
|  | Oct - Dec 2023 | 82 (2.3) | 326 (8.8) | 408 (5.6) | 206.2 ( 5.7) | 207.5 ( 5.6) |  |
| Scheme benefit level | Network/PMB | 1486 (41.3) | 1454 (39.2) | 2940 (40.2) | 1408.8 (38.9) | 1444.9 (39.2) | 0.011 |
|  | High day to day | 157 (4.4) | 193 (5.2) | 350 (4.8) | 189.7 ( 5.2) | 184.4 ( 5.0) |  |
|  | Low day to day | 1956 (54.3) | 2064 (55.6) | 4020 (55.0) | 2023.5 (55.9) | 2059.8 (55.8) |  |
| Initiation VL, copies/mL | 1000-9999 | 420 (11.7) | 362 (9.8) | 782 (10.7) | 382.5 (10.6) | 397.9 (10.8) | 0.008 |
|  | 10000-999999 | 1539 (42.8) | 1680 (45.3) | 3219 (44.0) | 1596.5 (44.1) | 1625.6 (44.1) |  |
|  | >999999 | 156 (4.3) | 279 (7.5) | 435 (6.0) | 217.1 ( 6.0) | 222.2 ( 6.0) |  |
|  | Missing | 1484 (41.2) | 1390 (37.5) | 2874 (39.3) | 1425.8 (39.4) | 1443.3 (39.1) |  |
| Initiation CD4 count, cells/μL | 0-49 | 203 (5.6) | 306 (8.2) | 509 (7.0) | 259.8 ( 7.2) | 259.2 ( 7.0) | 0.011 |
|  | 50-199 | 378 (10.5) | 481 (13.0) | 859 (11.8) | 417.7 (11.5) | 432.0 (11.7) |  |
|  | 200-349 | 381 (10.6) | 387 (10.4) | 768 (10.5) | 383.8 (10.6) | 383.3 (10.4) |  |
|  | 350-499 | 298 (8.3) | 311 (8.4) | 609 (8.3) | 297.5 ( 8.2) | 300.6 ( 8.1) |  |
| | ≥500 | 387 (10.8) | 418 (11.3) | 805 (11.0) | 405.1 (11.2) | 415.8 (11.3) | |
|  | Unknown | 1952 (54.2) | 1808 (48.7) | 3760 (51.4) | 1858.1 (51.3) | 1898.3 (51.5) |  |
|  |  | TEE (n = 3599, 49.2%) | TLD (n = 3711, 50.8%) | Total (n = 7310) | TEE (n=3622) | TLD (n=3689) | Standardised mean difference |
| TB at ART initiation | Yes (%) | 84 (2.3) | 114 (3.1) | 198 (2.7) | 104.5 ( 2.9) | 101.7 ( 2.8) | 0.008 |
| Pregnant at ART initiation | Yes (%) | 390 (10.8) | 198 (5.3) | 588 (8.0) | 281.8 ( 7.8) | 270.3 ( 7.3) | 0.017 |
| Hypercholesterolaemia | Yes (%) | 188 (5.2) | 216 (5.8) | 404 (5.5) | 201.7 ( 5.6) | 208.3 ( 5.6) | 0.003 |
| Hypertension | Yes (%) | 352 (9.8) | 374 (10.1) | 726 (9.9) | 363.7 (10.0) | 367.7 (10.0) | 0.003 |
| Diabetes mellitus | Yes (%) | 127 (3.5) | 96 (2.6) | 223 (3.1) | 111.7 ( 3.1) | 113.9 ( 3.1) | <0.001 |
| Chronic kidney disease | Yes (%) | 2 (0.1) | 3 (0.1) | 5 (0.1) | 2.1 ( 0.1) | 2.7 ( 0.1) | 0.006 |
| CVD risk factor* | Yes (%) | 500 (13.9) | 535 (14.4) | 1035 (14.2) | 679.2 (18.8) | 692.6 (18.8) | - |
| Statin use | Yes (%) | 81 (2.3) | 93 (2.5) | 174 (2.4) | 87.0 ( 2.4) | 90.2 ( 2.4) | 0.003 |
| Initiation BMI (including missing)** | Missing | 2856 (79.4) | 2461 (66.3) | 5317 (72.7) | 2720.3 (75.1) | 2590.6 (70.2) | 0.113 |
|  | Underweight/Normal | 225 (6.3) | 374 (10.1) | 599 (8.2) | 21.9 ( 0.6) | 20.7 ( 0.6) |  |
|  | Overweight | 244 (6.8) | 446 (12.0) | 690 (9.4) | 255.9 ( 7.1) | 305.2 ( 8.3) |  |
|  | Obese/ Severely obese | 274 (7.6) | 430 (11.6) | 704 (9.6) | 274.2 ( 7.6) | 323.7 ( 8.8) |  |

\*Composite of hypercholesterolaemia, hypertension, diabetes and chronic kidney disease
\*\*Not included in model to calculate IPTWs in main analysis

People were followed-up for a median of 21 months (IQR 10 to 33) until censoring, for a total of 12467 person-years. During follow up, 196 (5.3%) people who were initiated on TLD were changed to another dolutegravir-based regimen (n=84), or an efavirenz-based regimen (n=59) or another non-dolutegravir-based regimen (n=53), after a median of 212 days (IQR 90 to 426) from ART initiation. 636 (17.7%) people initiated on TEE were changed to TLD (n=536), another dolutegravir-based regimen (n=28), another efavirenz-based regimen (n=8) or another non-efavirenz based regimen (n=64), after a median of 306 days (IQR 125 to 634).

By the end of follow-up 4618 (63.2%) remained in care, 1765 (24.1%) had withdrawn, 739 (10.1%) experienced a gap in ART >6 months, 142 (1.9%) had died, and 46 (0.6%) had experienced a MACE. MACEs consisted of stroke (n=22), unstable angina (n=12), coronary revascularisation (n=8), and acute myocardial infarction (n=4). There were 18 MACEs with TLD after a median of 8 months (IQR 5 to 17), and 28 with TEE after a median of 4 months (IQR 3 to 13). The crude 3-year risk of MACE was 0.94% (95% CI 0.58 to 1.51) with TLD and 1.25% (0.85 to 1.83) with TEE.

#### Initiation cohort emulated target trial

In the emulated target trial intention to treat analysis, the standardised 3-year risk of MACE was 0.78% (0.37 to 1.32) with TLD and 0.96% (0.60 to 1.40) with TEE (risk ratio [RR] 0.81, 95% CI 0.35 to 1.60; risk difference [RD] −0.18 (−0.82 to 0.50). In the per-protocol analysis, the standardised 3-year risk of MACE was 0.62% (0.29 to 1.13) with TLD and 0.96% (0.58 to 1.40) with TEE (RR 0.65 [0.28 to 1.46]; RD −0.33% [−0.90 to 0.31]). In a sensitivity analysis including baseline BMI (with a category for missing) in the model to calculate IPTWs, there was no meaningful change in results (intention-to-treat analysis RR 0.81 [0.36 to 1.58], RD - 0.18% [−0.77 to 0.49]). In a sensitivity analysis with an interaction term between gender and treatment allocation in the MACE outcome model, there was no evidence of a difference in the effect of TLD on MACE in women (RR 0.76, 0.24 to 1.93), and in men (RR 0.86, 0.28 to 2.00, supplementary appendix Figure S1A).

### Transition cohort

In the transition cohort, we included 22,338 individuals who were receiving TEE in April 2020, and were potentially eligible for transition to TLD. Median (IQR) age was 41 (36-47) years and 61.7% were female (supplementary appendix Table S1). People were followed up for a median of 51 (25-51) months until censoring or database closure, for a total of 72514 person-years. By June 30, 2024 343 (1.5%) had died, 2546 (11.4%) had an ART gap >6 months, 6700 (30.0%) had withdrawn, 12494 (55.9%) remained in care without experiencing a MACE, and 255 (1.1%) had experienced a MACE. MACEs occurred after a median of 26 months (IQR 13 to 38) and included hospitalisation from stroke (n=109, 42.7%), unstable angina (n=102, 40.0%), acute myocardial infarction (n=24, 9.4%), coronary revascularisation (n=15, 5.9%) and peripheral arterial ischemia (n=3, 1.2%), and death from acute myocardial infarction (n=1, 0.4%) and stroke (n=1, 0.4%). During follow up, 2837 were transitioned to TLD while not viraemic after a median of 30 (IQR 15 to 39) months, and were included in the TLD arms of the emulated monthly sequential target trials. Those who were not viraemic and who remained on TEE at the same timepoint could be included in subsequent sequential trials, meaning individuals appeared in multiple trials.

#### Transition cohort emulated target trial

We therefore now describe the sequential trials’ population using person-trials. There were 2837 person-trials in the TLD arms and 706615 in the TEE arms of the 44 trials. The proportion of women (57.9% versus 62.2%), person-trials with CD4 count _≥_500 cells/µL (53.7% versus 59.0%), baseline cardiovascular risk factors (12.3% versus 15.9%) and missing BMI (66.1% versus 76.4%) were lower with TLD versus TEE (Table 3). However, distributions were similar among those with recorded BMI. Person-trials were slightly older (44 versus 43 years) and had been on ART for longer (7 versus 6 years) with TLD versus TEE. After IPTW, baseline characteristics were similar between groups (Table 3).

**Table 3:**
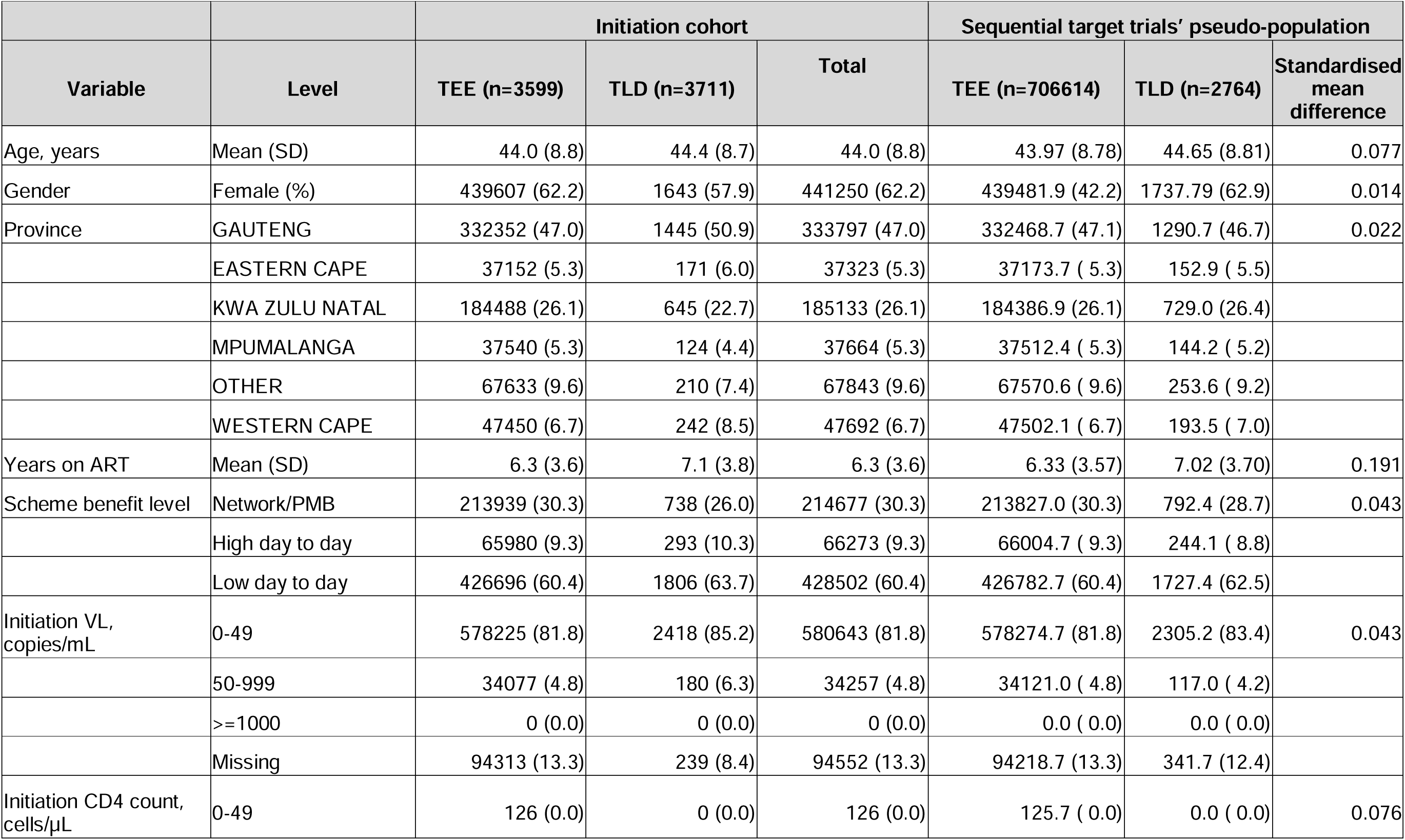
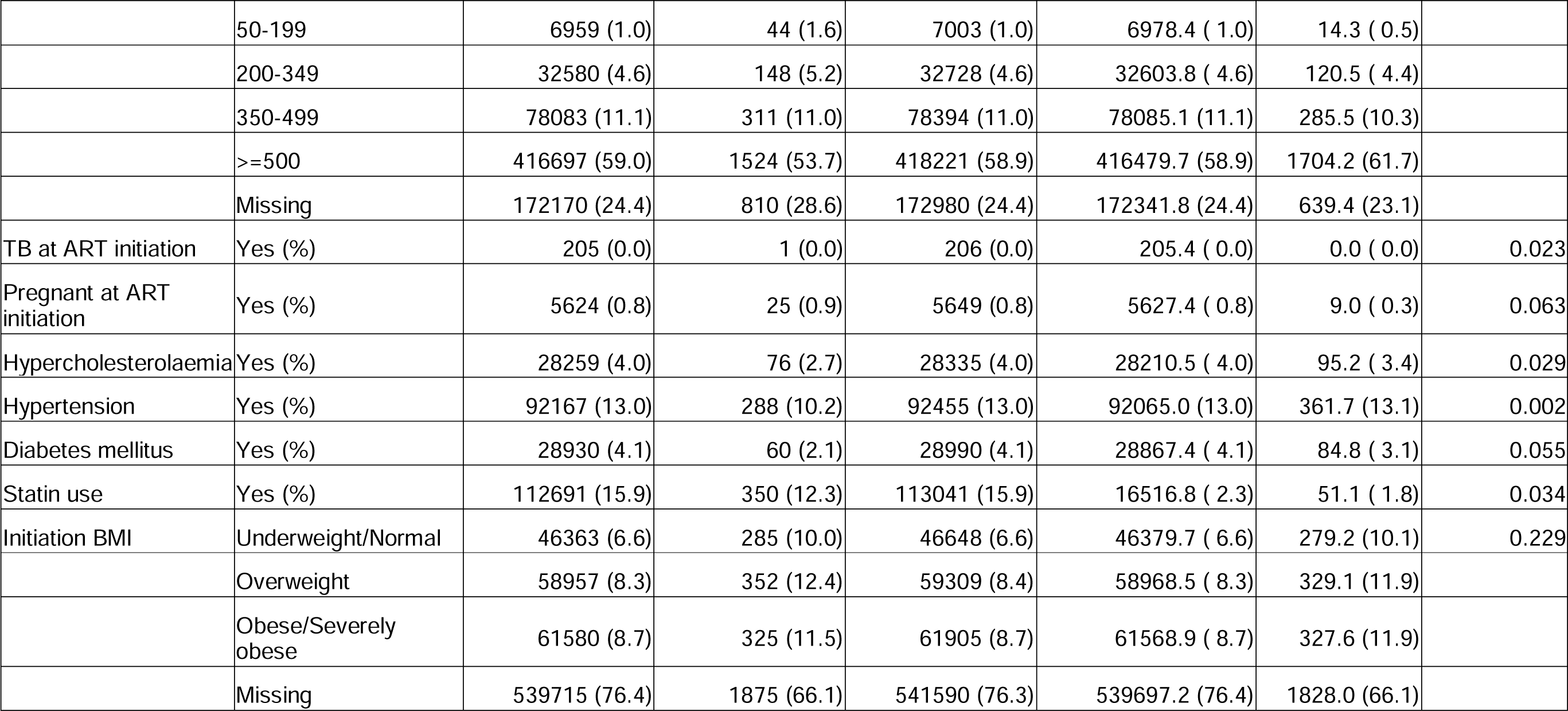
Baseline characteristics of transition cohort emulated target trial population, and pseudo-population after inverse probability of treatment weighting.

| Variable | Level | Initiation cohort |  |  | Sequential target trials' pseudo-population |  |  |
| --- | --- | --- | --- | --- | --- | --- | --- |
|  |  | TEE (n=3599) | TLD (n=3711) | Total | TEE (n=706614) | TLD (n=2764) | Standardised mean difference |
| Age, years | Mean (SD) | 44.0 (8.8) | 44.4 (8.7) | 44.0 (8.8) | 43.97 (8.78) | 44.65 (8.81) | 0.077 |
| Gender | Female (%) | 439607 (62.2) | 1643 (57.9) | 441250 (62.2) | 439481.9 (42.2) | 1737.79 (62.9) | 0.014 |
| Province | GAUTENG | 332352 (47.0) | 1445 (50.9) | 333797 (47.0) | 332468.7 (47.1) | 1290.7 (46.7) | 0.022 |
|  | EASTERN CAPE | 37152 (5.3) | 171 (6.0) | 37323 (5.3) | 37173.7 ( 5.3) | 152.9 ( 5.5) |  |
|  | KWA ZULU NATAL | 184488 (26.1) | 645 (22.7) | 185133 (26.1) | 184386.9 (26.1) | 729.0 (26.4) |  |
|  | MPUMALANGA | 37540 (5.3) | 124 (4.4) | 37664 (5.3) | 37512.4 ( 5.3) | 144.2 ( 5.2) |  |
|  | OTHER | 67633 (9.6) | 210 (7.4) | 67843 (9.6) | 67570.6 ( 9.6) | 253.6 ( 9.2) |  |
|  | WESTERN CAPE | 47450 (6.7) | 242 (8.5) | 47692 (6.7) | 47502.1 ( 6.7) | 193.5 ( 7.0) |  |
| Years on ART | Mean (SD) | 6.3 (3.6) | 7.1 (3.8) | 6.3 (3.6) | 6.33 (3.57) | 7.02 (3.70) | 0.191 |
| Scheme benefit level | Network/PMB | 213939 (30.3) | 738 (26.0) | 214677 (30.3) | 213827.0 (30.3) | 792.4 (28.7) | 0.043 |
|  | High day to day | 65980 (9.3) | 293 (10.3) | 66273 (9.3) | 66004.7 ( 9.3) | 244.1 ( 8.8) |  |
|  | Low day to day | 426696 (60.4) | 1806 (63.7) | 428502 (60.4) | 426782.7 (60.4) | 1727.4 (62.5) |  |
| Initiation VL, copies/mL | 0-49 | 578225 (81.8) | 2418 (85.2) | 580643 (81.8) | 578274.7 (81.8) | 2305.2 (83.4) | 0.043 |
|  | 50-999 | 34077 (4.8) | 180 (6.3) | 34257 (4.8) | 34121.0 ( 4.8) | 117.0 ( 4.2) |  |
|  | >=1000 | 0 (0.0) | 0 (0.0) | 0 (0.0) | 0.0 ( 0.0) | 0.0 ( 0.0) |  |
|  | Missing | 94313 (13.3) | 239 (8.4) | 94552 (13.3) | 94218.7 (13.3) | 341.7 (12.4) |  |
| Initiation CD4 count, cells/μL | 0-49 | 126 (0.0) | 0 (0.0) | 126 (0.0) | 125.7 ( 0.0) | 0.0 ( 0.0) | 0.076 |
|  |  | TEE (n=3599) | TLD (n=3711) | Total | TEE (n=706614) | TLD (n=2764) | Standardised mean difference |
|  | 50-199 | 6959 (1.0) | 44 (1.6) | 7003 (1.0) | 6978.4 ( 1.0) | 14.3 ( 0.5) |  |
|  | 200-349 | 32580 (4.6) | 148 (5.2) | 32728 (4.6) | 32603.8 ( 4.6) | 120.5 ( 4.4) |  |
|  | 350-499 | 78083 (11.1) | 311 (11.0) | 78394 (11.0) | 78085.1 (11.1) | 285.5 (10.3) |  |
|  | >=500 | 416697 (59.0) | 1524 (53.7) | 418221 (58.9) | 416479.7 (58.9) | 1704.2 (61.7) |  |
|  | Missing | 172170 (24.4) | 810 (28.6) | 172980 (24.4) | 172341.8 (24.4) | 639.4 (23.1) |  |
| TB at ART initiation | Yes (%) | 205 (0.0) | 1 (0.0) | 206 (0.0) | 205.4 ( 0.0) | 0.0 ( 0.0) | 0.023 |
| Pregnant at ART initiation | Yes (%) | 5624 (0.8) | 25 (0.9) | 5649 (0.8) | 5627.4 ( 0.8) | 9.0 ( 0.3) | 0.063 |
| Hypercholesterolaemia | Yes (%) | 28259 (4.0) | 76 (2.7) | 28335 (4.0) | 28210.5 ( 4.0) | 95.2 ( 3.4) | 0.029 |
| Hypertension | Yes (%) | 92167 (13.0) | 288 (10.2) | 92455 (13.0) | 92065.0 (13.0) | 361.7 (13.1) | 0.002 |
| Diabetes mellitus | Yes (%) | 28930 (4.1) | 60 (2.1) | 28990 (4.1) | 28867.4 ( 4.1) | 84.8 ( 3.1) | 0.055 |
| Statin use | Yes (%) | 112691 (15.9) | 350 (12.3) | 113041 (15.9) | 16516.8 ( 2.3) | 51.1 ( 1.8) | 0.034 |
| Initiation BMI | Underweight/Normal | 46363 (6.6) | 285 (10.0) | 46648 (6.6) | 46379.7 ( 6.6) | 279.2 (10.1) | 0.229 |
|  | Overweight | 58957 (8.3) | 352 (12.4) | 59309 (8.4) | 58968.5 ( 8.3) | 329.1 (11.9) |  |
|  | Obese/Severely obese | 61580 (8.7) | 325 (11.5) | 61905 (8.7) | 61568.9 ( 8.7) | 327.6 (11.9) |  |
|  | Missing | 539715 (76.4) | 1875 (66.1) | 541590 (76.3) | 539697.2 (76.4) | 1828.0 (66.1) |  |

**Table 4:** Observed and standardised 3-year risk of major adverse cardiovascular events with TLD versus TEE in the initiation and transition cohort emulated target trials.

|  | N | Median fu time, years (IQR) | Person-years fu | Events | Crude 36-month risk | Standardised 36-month risk | Risk difference | Risk ratio |
| --- | --- | --- | --- | --- | --- | --- | --- | --- |
| Initiation cohort intention to treat analysis |  |  |  |  |  |  |  |  |
| TLD | 3,711 | 1.42 (0.83 to 2.25) | 5,746 | 18 | 0.94% (0.58 to 1.51) | 0.78% (0.37 to 1.32) | -0.18% (-0.82 to 0.50) | 0.81 (0.35 to 1.59) |
| TEE | 3,599 | 1.92 (0.92 to 3.00) | 6,721 | 28 | 1.25% (0.85 to 1.83) | 0.96% (0.60 to 1.40) |  |  |
| Initiation cohort per-protocol analysis |  |  |  |  |  |  |  |  |
| TLD | 3,711 | 1.33 (0.75 to 2.17) | 5,538 | 16 | 0.87% (0.51 to 1.44) | 0.62% (0.29 to 1.13) | -0.33% (-0.90 to 0.31) | 0.65 (0.28 to 1.46) |
| TEE | 3,599 | 1.58 (0.75 to 2.83) | 6,014 | 26 | 1.30% (0.87 to 1.92) | 0.96% (0.58 to 1.40) |  |  |
| Transition cohort intention to treat analysis |  |  |  |  |  |  |  |  |
| TLD | 2,837 | 1.42 (0.83 to 2.50) | 4,642 | 19 | 1.23% (0.76 to 1.95) | 1.09% (0.48 to 1.99) | -0.12% (-0.75 to 0.75) | 0.90 (0.41 to 1.64) |
| TEE | 706,615* | 2.08 (1.17 to 3.00) | 1,391,382** | 5420 | 1.17% (1.14 to 1.20) | 1.21% (1.05 to 1.41) |  |  |
| Transition cohort per-protocol analysis |  |  |  |  |  |  |  |  |
| TLD | 2,837 | 1.33 (0.75 to 2.33) | 4,315 | 13 | 0.90% (0.50 to 1.58) | 0.75% (0.28 to 1.48) | -0.46% (-0.95 to 0.33) | 0.62 (0.24 to 1.28) |
| TEE | 706,615* | 1.92 (1.00 to 2.92) | 1,308,491 | 4834 | 1.11% (1.08 to 1.14) | 1.20% (1.01 to 1.41) |  |  |

Within the sequential trials, person-trials were followed for a median of 25 (14 to 36) months, for a total of 1396025 person-trial-years. Among those who were allocated to TLD at trial baseline, 302 (10.6%) subsequently changed regimen to an efavirenz-based regimen (n=202), another dolutegravir-based regimen (n=65) or another non-dolutegravir-based regimen (n=35), after a median of 8 (4-13) months. Of those who continued TEE at baseline, 81326 (11.5%) subsequently changed regimen, to TLD (n=72701), another dolutegravir-based regimen (n=2545), another efavirenz-based regimen (n=1586) or another non-efavirenz based regimen (n=4494), after a median of 14 (7-24) months. There were 19 MACEs with TLD after a median of 9 months (IQR 6.5 to 15), and 5420 with TEE after a median of 15 months (IQR 7 to 23). The crude 3-year risk of MACE with TLD was 1.23 (0.76 to 1.95) and 1.16 (1.13 to 1.19) with TEE.

In the sequential trials intention-to-treat analysis, the standardised 3-year risk of MACE was 1.09% (0.48 to 1.99) with TLD and 1.21% (1.05 to 1.41) with TEE (RR 0.90, [0.41 to 1.64]; RD −0.12% [−0.75 to 0.75]). In the per-protocol analysis, the standardised 3-year risk of MACE was 0.75% (0.28 to 1.48) with TLD and 1.20% (1.01 to 1.41) with TEE (RR 0.62, [0.24 to 1.28]; RD −0.46 [−0.95 to 0.33]). In a sensitivity analysis including baseline BMI (with a category for missing) in the model to calculate IPTWs, there was no meaningful change in results (intention-to-treat analysis RR 1.03 [0.49-1.86], RD −0.03% [−0.60-1.04]). In a sensitivity analysis with an interaction term between gender and treatment allocation in the MACE outcome model, there was no evidence of a difference in the effect of TLD on MACE in women (RR 1.14, 0.30 to 2.53), and in men (RR 0.72, 0.28 to 1.28, supplementary appendix Figure S1B).

## DISCUSSION

In this large South African cohort of PLHIV, newly initiating and already receiving ART, we found no evidence of an increased risk of MACE over three years among people taking TLD compared to TEE.

Our findings align with most previous studies from high-income settings, which have generally not found evidence of increased MACE risk with INSTIs. A systematic review of nine clinical trials with 6647 person-years of follow-up found no evidence of increased serious adverse cardiovascular events with dolutegravir (15/2202, 0.7%) versus other antiretrovirals (8/2215, 0.4%, relative risk 1.69, 95% CI 0.71 to 4.03), although numbers were small and study design and comparator antiretrovirals were heterogenous.^22^ Two retrospective cohort studies using North American health insurance data and IPTW among 20242^9^ and 14076^10^ new ART initiators found that INSTIs were associated with lower (HR 0.79, 0.64 to 0.96)^9^ or similar (HR 1.30, 0.95 to 1.88) risk of MACE in the first year or two of follow-up. In contrast, a retrospective cohort study of 29340 individuals in European and Australian cohorts found that INSTI use was associated with an almost two times higher risk of MACE in people with 0 to 6 months of cumulative INSTI exposure (incidence rate ratio [IRR] 1·85, 1·44 to 2·39), which remained elevated up to 24 months of INSTI exposure (IRR 1·46, 1·13 to 1·88), before equalising between 24 to 36 months (IRR 0·89, 0·62–1·29).^4^ While the study design has been questioned,^8^ these findings have raised concern for global HIV treatment programmes, and resulted in calls for further studies.^23^ Subsequent target trial emulations in North American and European cohorts (n=87990 individuals) found similar risks of MACE among people newly initiating INSTI versus non-INSTI ART (4-year aRD 0·01%, −0·43 to 0·36) and among people transitioned to INSTI versus remaining on non-INSTI ART (aRD −0·07%, −0·60 to 0·52),^8^ while an emulated target trial using data from 5362 Swiss people found no difference in MACE between those initiating INSTIs versus non-INSTIs (adjusted hazard ratio 0.80, 0.46 to 1.39)^7^.

Our study adds substantially to the evidence base by specifically evaluating the risk of MACE with TLD in an LMIC setting, where this regimen is most widely used. Furthermore, we include people transitioning from TEE to TLD, who make up the majority of people exposed to TLD globally, and directly address the causal question of whether this transition increases MACE risk. Further strengths of our study include the use of comprehensive, longitudinal health insurance claims data that include both primary care treatment and cardiovascular risk factor data, and secondary care and vital statistics data on cardiovascular hospitalisations and deaths. While such health insurance datasets have been used for research in high income settings, their use in LMICs is more recent,^24,25^ providing an opportunity in settings where large research cohorts with long follow-up time have not yet been established. A potential weakness of using health insurance data^24^ is that our results may not be generalisable to public sector settings, where most people receive HIV care in South Africa.^26^ Nevertheless, in private sector settings, stroke diagnosis may be more reliable than in the public sector, where there is often limited access to computed tomography or magnetic resonance imaging scans^27^. We used an emulated target trial methodology to minimise bias in this analysis of observational data. However, we cannot rule out residual confounding, in particular by BMI, which was missing for many participants. Clinicians may have avoided using TLD in people with a high BMI, meaning that the TLD group would have had a lower baseline BMI, which would lower their baseline cardiovascular risk. However, to calculate the IPTW we included baseline hypertension, diabetes and hypercholesterolaemia status, which are on the pathway between BMI and cardiovascular risk. While our study is one of the largest in an LMIC to evaluate cardiovascular outcomes among people on ART, with over 6500 individuals receiving TLD, the upper bound of the 95% confidence interval for the risk difference in the transition cohort was 0.75%, meaning we cannot rule out an extra 7.5 MACEs per 1000 individuals with TLD over 3 years. Larger studies with greater follow-up time are therefore required to determine if TLD associated weight gain translates to higher MACE risk beyond three years.

Overall, our results provide reassurance that for the over 20 million people in LMICs who are now using TLD instead of TEE, there is no large increase in MACE risk in the short to medium term. Given the benefits of TLD in terms of viral suppression and current low prevalence of resistance, these findings support the ongoing use of TLD as recommended by WHO.

## Data Availability

The data used for this analysis cannot be shared publicly because of the legal (Protection of Personal Information Act) and ethical requirements regarding the use of routinely collected clinical data in South Africa, and because our approved study protocol does not include permission to share the data. Researchers may request access to the data from Discovery Health (contact details obtainable upon request to corresponding author).

## ABBREVIATIONS

ART: Antiretroviral therapy
BMI: Body-mass index
INSTI: Integrase strand transferase inhibitor
IPTWs: Inverse probability of treatment weights
IPCWs: Inverse probability of censoring weights
LMIC: Low- and middle-income country
MACE: Major adverse cardiovascular event
PLHIV: People living with HIV
RR: Risk ratio
RD: Risk difference
TEE: Tenofovir disoproxil fumarate, emtricitabine and efavirenz
TLD: Tenofovir disoproxil fumarate, lamivudine and dolutegravir
WHO: World Health Organization

## AUTHORSHIP CONTRIBUTIONS

JD, NG and SC conceptualized the study. JD, LL, KT, SC and NG managed the project. XM, CP, DJ and SC oversaw data collection. XM, CP, DJ, SC and JvdM oversaw data curation. XM, CP, JvdM and JD have directly accessed and verified the underlying data. JD, LL, JvdM, KA, JAB and CB analysed the data. JD drafted the manuscript. JD and NG had full access to all the data in the study and had final responsibility for the decision to submit for publication. All authors contributed to interpretation of results, critically reviewed and edited the manuscript, and consented to final publication.

## ACKNOWLEDGEMENTS

We would like to acknowledge the staff and patients in the Discovery Health scheme.

---

**Figure 1:**
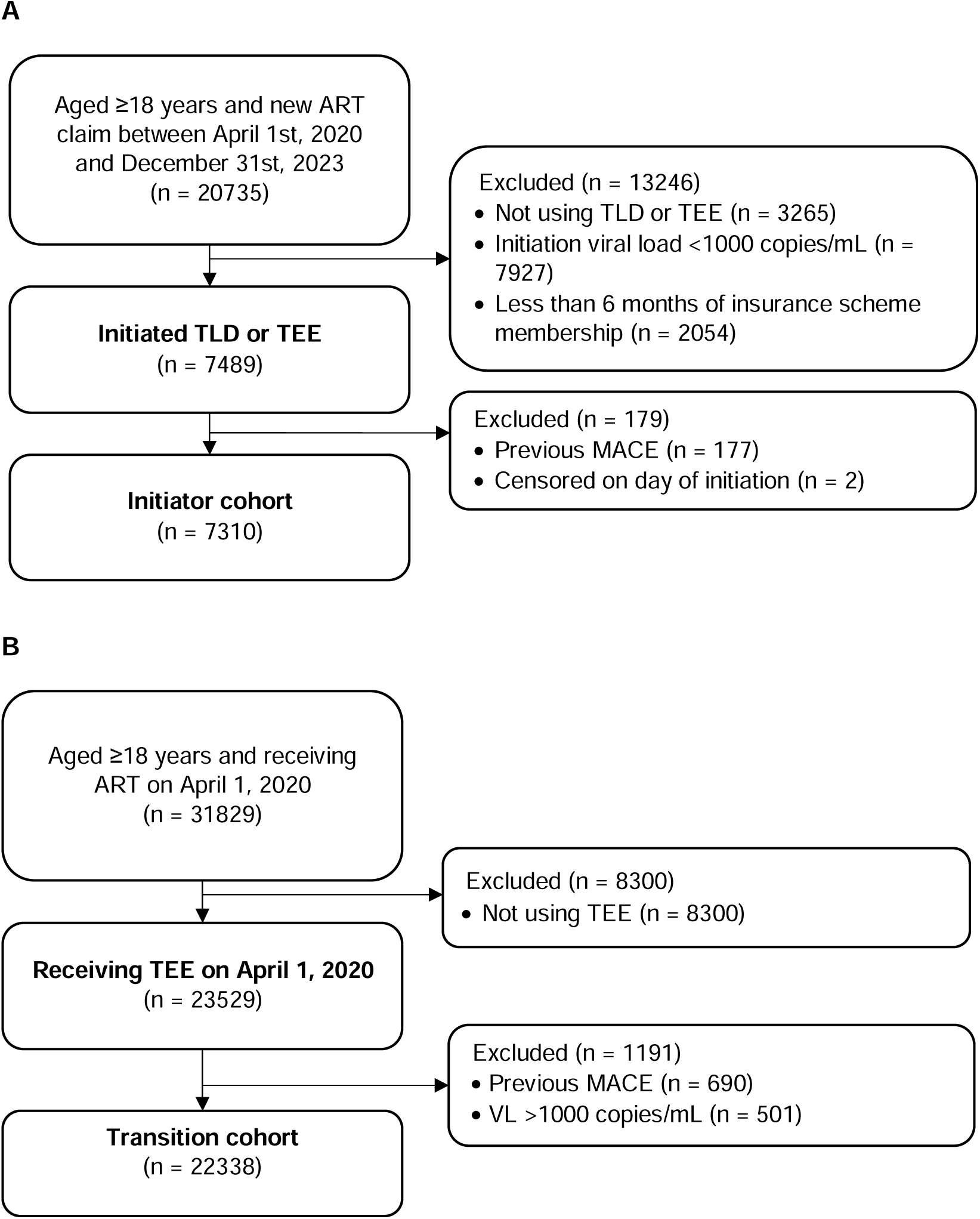

