## supplementary appendix for "Risk of major adverse cardiovascular events with dolutegravir versus efavirenz-based antiretroviral therapy: emulated target trials using routine, de-identified data from South Africa"

### 1. Figure S1 Forest plot of overall and sub-group effects by gender of TLD versus TEE on the risk of MACE in the initiation and transition emulated target trials

A) Initiation cohort

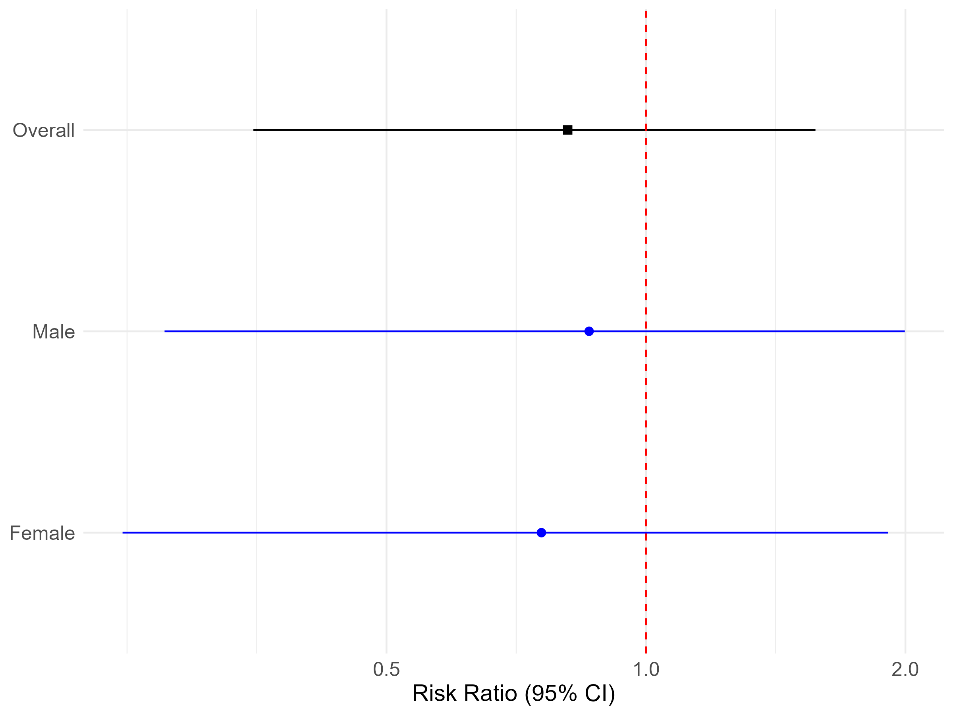

B) Transition cohort

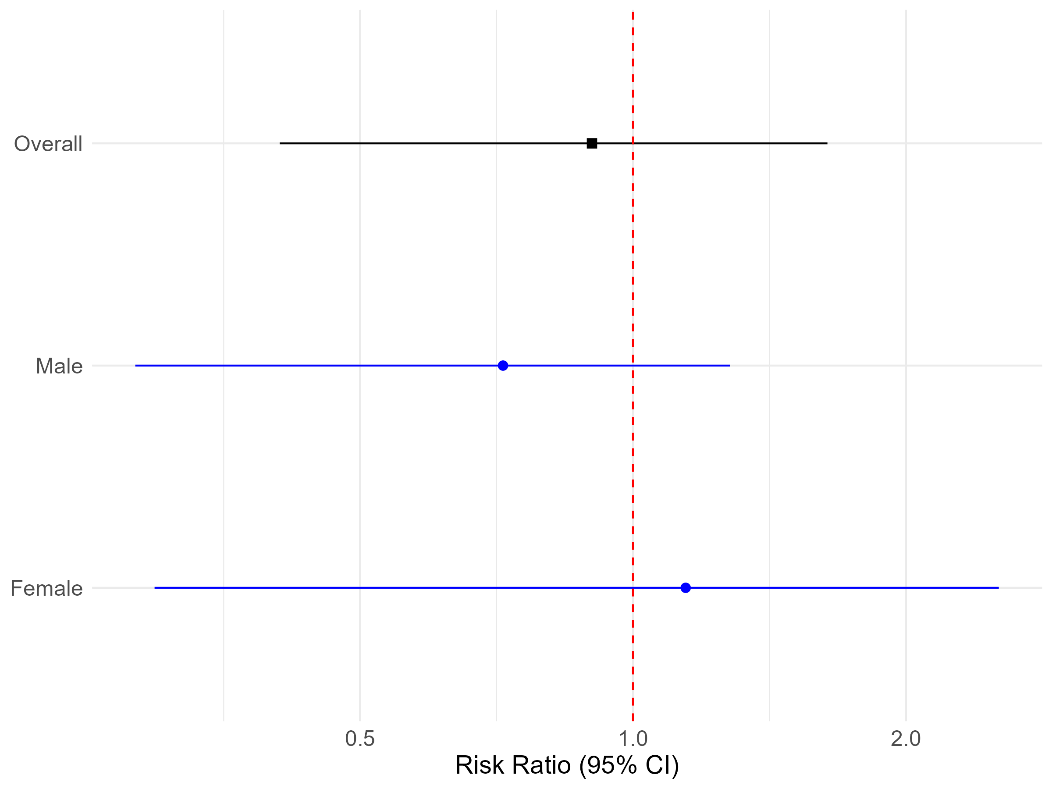

### 2. Table S1 Characteristics of individuals included in the transition cohort, at the start of the cohort on April 1^st^, 2020 (n = 22338)

| **Variable** | **Levels** | **Total** |
| --- | --- | --- |
| Age (years) | Median (IQR) | 41.0 (36.0 to 47.0) |
| Gender | F | 13788 (61.7) |
|  | M | 8550 (38.3) |
| Province | GAUTENG | 10563 (47.3) |
|  | EASTERN CAPE | 1185 (5.3) |
|  | KWA ZULU NATAL | 5713 (25.6) |
|  | MPUMALANGA | 1170 (5.2) |
|  | OTHER | 2181 (9.8) |
|  | WESTERN CAPE | 1526 (6.8) |
| Time since ART initiation, years | Median (IQR) | 4.4 (2.3 to 7.5) |
| Scheme benefit level | Network/PMB | 7032 (31.5) |
|  | High day to day | 1990 (8.9) |
|  | Low day to day | 13316 (59.6) |
| Baseline VL (copies/mL) | 0-49 | 16573 (74.2) |
|  | 50-999 | 1156 (5.2) |
|  | >=1000 | 310 (1.4) |
|  | Missing | 4299 (19.2) |
| Time since baseline VL (days) | Median (IQR) | -76.0 (-147.0 to -27.0) |
| Baseline CD4 count (cells/µL) | 0-200 | 355 (1.6) |
|  | 201-349 | 1187 (5.3) |
|  | 350-499 | 2431 (10.9) |
|  | >=500 | 11501 (51.5) |
|  | Unknown | 6864 (30.7) |
| Time since baseline CD4 count (days) | Median (IQR) | -79.0 (-154.0 to -29.0) |
| TB at baseline | No | 22246 (99.6) |
|  | Yes | 92 (0.4) |
| Pregnant at ART baseline | NOT PREGNANT | 22036 (98.6) |
|  | PREGNANT | 302 (1.4) |
| Hypercholesterolaemia | No | 21633 (96.8) |
|  | Yes | 705 (3.2) |
| Hypertension | No | 19803 (88.7) |
|  | Yes | 2535 (11.3) |
| Diabetes mellitus | No | 21539 (96.4) |
|  | Yes | 799 (3.6) |
| Chronic kidney disease | No | 22327 (100.0) |
|  | Yes | 11 (0.0) |
| CVD risk factor | No | 19236 (86.1) |
|  | Yes | 3102 (13.9) |
| Statin use | No | 21626 (96.8) |
|  | Yes | 712 (3.2) |
| BMI (inc missing) | Missing | 18437 (82.5) |
|  | Underweight/Normal | 1153 (5.2) |
|  | Overweight | 1391 (6.2) |
|  | Obese/Severely obese | 1357 (6.1) |
| BMI | Underweight/Normal | 1153 (29.6) |
|  | Overweight | 1391 (35.7) |
|  | Obese/Severely obese | 1357 (34.8) |
| Time since baseline BMI (days) | Median (IQR) | -781.0 (-1672.0 to -326.0) |

### 3. R code for initiation cohort emulated target trial analysis

library(here); library(magrittr);library(tidyverse); library(janitor);

library(ggplot2); library (margins); library(tableone)

library(survey); library(kableExtra); library(splines)

\### Load the dataset

here()

\## Main dataset including follow up time

\# Data structured outlined here: https://journals.sagepub.com/doi/10.1177/1536867X0400400403

\# One row per month per participant, time varying covariates updated monthly

fu_data_exp_itt <- readRDS(file = here("analysis_datasets", "fu_data_exp_itt.rds"))

\## Data at baseline

\# One row per participant, covariates as at baseline

base_df <- readRDS(file = here("analysis_datasets", "base_df.rds"))

\### Set seed for reproducibility

set.seed(123)

\### Calculate the propensity for treatment

\# Estimate the propensity score, weights and stabilised weights using baseline data only

ps_fit <- glm(dtg ~ gender + init_age + province_gp + benefit_gp + time_cat +

init_cd4_cat + init_vl_cat + init_tb + init_preg + init_dm + init_htn + init_hypchol + init_statin, data = base_df,

family = quasibinomial)

\# Add propensity score to the base dataset

base_df$ps <- fitted(ps_fit)

\# Plot ps for dtg versus non-dtg

ggplot(base_df, aes(x = ps, fill = factor(dtg))) +

geom_histogram(position = "identity", alpha = 0.5, bins = 30) +

labs(title = "Distribution of ps by dtg Status",

x = "pr",

y = "Count",

fill = "dtg") +

theme_minimal()

\### calculate the INVERSE PROBABILITY of treatment weights

\# (1/prob of treatment in treated, 1/prob of not being treated in untreated)

base_df$ps_w <- with(base_df, dtg/ps + (1-dtg)/(1-ps))

\## calculate the STABILISED INVERSE PROBABILITY of treatment weights

\# (prop exposed/prob of treatment in treated, prop unexposed/prob of not being treated in untreated)

base_df$ps_w_stab <- with(base_df,

ifelse(dtg == 1,

mean(dtg == 1) / ps,

mean(dtg == 0) / (1 - ps)))

\# Add the propensity score and weights to the follow up dataset

fu_data_exp_itt <- base_df %>%

select(study_id, ps, ps_w, ps_w_stab) %>%

right_join(fu_data_exp_itt, by = "study_id")

\### Creat table to check if weighted pseudo-population is balanced

survey_design <- svydesign(ids = ~1, data = base_df, weights = ~ps_w_stab)

table <- svyCreateTableOne(

vars = c("init_age", "gender", "province_gp", "time_cat", "benefit_gp", #"init_scheme_dur",

"init_vl_cat", "init_cd4_cat", "init_tb", "init_preg", "init_hypchol", "init_htn",

"init_dm", "init_ckd", "init_statin", "bmi_cat_miss2"), # Covariates to be included

strata = "dtg", # Group by treatment status

data = survey_design	# Apply IPTW weights

)

print(table, smd = T)

\### Use the weights in the outcome model

\# make a new dtg exposure variable so that when changing the exposure as part of standardization, the original treatment allocations unaffected

fu_data_exp_itt %<>%

mutate(dtg_gitt = dtg)

#Fit the outcome model with IPTW (stabilised) # Note no meaningful difference in results when using splines, therefore not used

fit_itt <- glm(cvd ~ dtg_gitt + cummonth + cummonth^2 + dtg_gitt:cummonth + dtg_gitt:cummonth^2, data = fu_data_exp_itt,

family = quasibinomial, weights = ps_w_stab)

#Estimate potential outcomes under treatment

fu_data_exp_itt$dtg_gitt <- 1

fu_data_exp_itt$pred1_itt <- predict(fit_itt, newdata = fu_data_exp_itt, type = "response")

#Estimate potential outcomes under control

fu_data_exp_itt$dtg_gitt <- 0

fu_data_exp_itt$pred0_itt <- predict(fit_itt, newdata = fu_data_exp_itt, type = "response")

\### Calculate standardised 3 year risk ratio & risk difference, allowing monthly probability to vary

\# make vector of max_fu month probs under treatment

pred1_max_fum_pr <- fu_data_exp_itt %>%

data.frame() %>%

select(cummonth, pred1_itt) %>%

distinct() %>%

pull(pred1_itt)

\# make vector of max_fu month probs under control

pred0_max_fum_pr <- fu_data_exp_itt %>%

data.frame() %>%

select(cummonth, pred0_itt) %>%

distinct() %>%

pull(pred0_itt)

#Compute standardised risk under treatment

std_rx1_var <- 1- (prod(1- pred1_max_fum_pr))

std_rx1_var

#Compute standardised 3 year risk under treatment

std_rx0_var <- 1- (prod(1- pred0_max_fum_pr))

std_rx0_var

#Compute standardised risk difference

std_risk_difference_var <- std_rx1_var - std_rx0_var

std_risk_difference_var

#Compute standardised risk ratio

std_risk_ratio_var <- std_rx1_var/std_rx0_var

std_risk_ratio_var

\### BOOTSTRAP to estimate CIs in ITT

\# make a new dtg exposure variable so that when changing the exposure as part of standardisation, the original treatment allocations unaffected

fu_data_exp_itt %<>%

mutate(dtg_gitt_boot = dtg)

output <- data.frame(

boot = numeric(0),

std_rx1_var_boot = numeric(0),

std_rx0_var_boot = numeric(0),

std_risk_difference_var_boot = numeric(0),

std_risk_ratio_var_boot = numeric(0),

)

boots <- c(1:500)

for(i in boots){

\## Sample study_ids with replacement

in_study_ids <- slice_sample(distinct(fu_data_exp_itt, study_id), prop = 1, replace = T)

\# set unique identifiers for replicated study_id's

in_study_ids_rep <- in_study_ids %>% # vector of study_id's as a result of sampling study_id's with replacement

group_by(study_id) %>% #filter(n()>1)

mutate(rep_study_id = paste(row_number(), study_id, sep = "_")) %>% #filter(n()>1) %>% arrange(study_id, rep_study_id) %>%

ungroup()

\# get baseline dataset for the sampled study ids

in_base_data <- in_study_ids_rep %>%

left_join(base_df, by = "study_id")

\# create fu data_set data for sampled study ids

in_data <- in_study_ids_rep %>%

left_join(fu_data_exp_itt, by = "study_id", relationship = "many-to-many")

#Estimate the PS (use baseline data)

ps_fit_boot <- glm(dtg ~ gender + init_age + province_gp + benefit_gp + time_cat + #+ init_scheme_dur

init_cd4_cat + init_vl_cat + init_tb + init_preg + init_dm + init_htn + init_hypchol + init_statin , data = in_base_data,

family = quasibinomial)

\# add it to the dataset

in_base_data$ps_bt <- fitted(ps_fit_boot)

\# calculate the IPTW

in_base_data$ps_bt_w <- with(in_base_data, dtg/ps_bt + (1-dtg)/(1-ps_bt))

\# calculate the STABILISED INVERSE PROBABILITY of treatment weights (prop exposed/prob of treatment in treated, prop unexposed/prob of not being treated in untreated)

in_base_data$ps_bt_w_stab <- with(in_base_data,

ifelse(dtg==1,

mean(dtg==1)/ps_bt,

mean(dtg==0)/(1-ps_bt)))

in_data <- in_base_data %>%

select(rep_study_id, ps_bt, ps_bt_w, ps_bt_w_stab) %>%

right_join(in_data, by = "rep_study_id")

\# run the model weighted with the iptw

model_sample <- glm(cvd ~ dtg_gitt_boot + cummonth + cummonth^2 + dtg_gitt_boot:cummonth + dtg_gitt_boot:cummonth^2

, data = in_data, family = quasibinomial, weights = ps_bt_w_stab)

#Estimate potential outcomes under treatment

in_data$dtg_gitt_boot <- 1

in_data$pred1_itt <- predict(model_sample, newdata = in_data, type = "response")

#Estimate potential outcomes under control

in_data$dtg_gitt_boot <- 0

in_data$pred0_itt <- predict(model_sample, newdata = in_data, type = "response")

\# make vector of max_fu month probs under treatment

pred1_max_fum_pr_boot <- in_data %>%

data.frame() %>%

select(cummonth, pred1_itt) %>%

distinct() %>%

pull(pred1_itt)

\# make vector of max_fu month probs under control

pred0_max_fum_pr_boot <- in_data %>%

data.frame() %>%

select(cummonth, pred0_itt) %>%

distinct() %>%

pull(pred0_itt)

#Compute standardised 3 year risk under treatment

std_rx1_var_boot <- 1- (prod(1- pred1_max_fum_pr_boot))

std_rx1_var_boot

#Compute standardised 3 year risk under control

std_rx0_var_boot <- 1- (prod(1- pred0_max_fum_pr_boot))

std_rx0_var_boot

#Compute standardised risk difference

std_risk_difference_var_boot <- std_rx1_var_boot - std_rx0_var_boot

std_risk_difference_var_boot

#Compute standardised risk ratio

std_risk_ratio_var_boot <- std_rx1_var_boot/std_rx0_var_boot

std_risk_ratio_var_boot

\# all outputs combine in one tbl

boot_output <- data.frame(

boot = i,

std_rx1_var_boot = std_rx1_var_boot,

std_rx0_var_boot = std_rx0_var_boot,

std_risk_difference_var_boot = std_risk_difference_var_boot,

std_risk_ratio_var_boot = std_risk_ratio_var_boot,

)

output <- rbind(output, boot_output)

}

\# save the output to avoid regenerating

saveRDS(output, file = here("output.rds"))

output <- readRDS(file = here("output.rds"))

\# Calculate the 95% confidence interval of std risk under treatment using percentiles

std_rx1_ci_lower <- quantile(output$std_rx1_var_boot, 0.025)

std_rx1_ci_lower

std_rx1_ci_upper <- quantile(output$std_rx1_var_boot, 0.975)

std_rx1_ci_upper

\# Calculate the 95% confidence interval of std risk under control

std_rx0_ci_lower <- quantile(output$std_rx0_var_boot, 0.025)

std_rx0_ci_lower

std_rx0_ci_upper <- quantile(output$std_rx0_var_boot, 0.975)

std_rx0_ci_upper

\# Calculate the 95% confidence interval of std risk difference using percentiles

std_rd_ci_lower <- quantile(output$std_risk_difference_var_boot, 0.025)

std_rd_ci_lower

std_rd_ci_upper <- quantile(output$std_risk_difference_var_boot, 0.975)

std_rd_ci_upper

\# Calculate the 95% confidence interval of std risk ratio

std_rr_ci_lower <- quantile(output$std_risk_ratio_var_boot, 0.025)

std_rr_ci_lower

std_rr_ci_upper <- quantile(output$std_risk_ratio_var_boot, 0.975)

std_rr_ci_upper

\### Per protocol analysis with censoring when change ART, so weight for censoring

\# Notes for censoring also taken from here: https://journals.sagepub.com/doi/10.1177/1536867X0400400403

\# load data

fu_data_exp_pp <- readRDS(file = here("analysis_datasets", "fu_data_exp_pp.rds"))

\# make a new dtg exposure variable so that when changing the exposure as part of standardization, the original treatment allocations unaffected

fu_data_exp_pp %<>%

mutate(dtg_gpp = dtg)

\### n = 5078 same as ITT so propensity of treatment will be the same as calculated at baseline in ITT analysis, so add this and unstabilised weight to fu dataframe

fu_data_exp_pp <- base_df %>%

select(study_id, ps, ps_w, ps_w_stab) %>%

right_join(fu_data_exp_pp, by = "study_id")

\### Logistic regression model to generate probability of censoring

pr_cens <- glm(cens_pp ~ init_art + gender + init_age + province_gp + benefit_gp + time_cat +

init_cd4_cat + init_vl_cat + init_tb + init_preg + init_dm + init_htn +

init_hypchol + init_statin + vl_cat + cd4_cat + preg + tb + dm + chol + statin +

cummonth, data = fu_data_exp_pp

, family = quasibinomial)

\# generate the probabilities and bind to the dataset

pr_cens_df <- data.frame(predict(pr_cens, type = "response"))

pr_cens_df %<>%

rename(pr_cens = 1)

fu_data_exp_pp %<>%

bind_cols(pr_cens_df)

\# We have now estimated the probability of each person being censored in each month,

\# given their covariate history. Subtracting these estimated probabilities from 1 results

\# in an estimate of the probability of a person being uncensored in each month.

fu_data_exp_pp %<>%

mutate(pr_cens = 1 - pr_cens)

\# To derive each subject’s estimated probability of their complete censoring history up to

\# each month, we multiply the estimated probabilities of being

\# uncensored for each month cumulatively over time.

fu_data_exp_pp %<>%

arrange(study_id, cummonth) %>%

group_by(study_id) %>%

mutate(pr_cens_cum = cumprod(pr_cens)) %>%

ungroup()

\# Add unstabilised weights: 1/pr_cens_cum

fu_data_exp_pp %<>%

mutate(pr_cens_wt = 1/pr_cens_cum)

summary(fu_data_exp_pp$pr_cens_wt)

ggplot(fu_data_exp_pp, aes(x = pr_cens_wt)) +

geom_histogram(binwidth = 0.005, fill = "skyblue", color = "black") +

labs(title = "Histogram of unstabilised pr_cens_wt", x = "pr_cens_wt", y = "Frequency")

\# add stabilised weights

\### Logistic regression model to generate probability of censoring but without time varying covariates

pr_cens <- glm(cens_pp ~ init_art + gender + init_age + province_gp + benefit_gp + time_cat +

init_cd4_cat + init_vl_cat + init_tb + init_preg + init_dm + init_htn +

init_hypchol + init_statin + cummonth,

data = fu_data_exp_pp,

family = quasibinomial)

\# generate the probabilities and bind to the dataset

pr_cens_df <- data.frame(predict(pr_cens, type = "response"))

pr_cens_df %<>%

rename(pr_cens_stab = 1)

fu_data_exp_pp %<>%

bind_cols(pr_cens_df)

\# We have now estimated the probability of each person being censored in each month,

\# given their covariate history. Subtracting these estimated probabilities from 1 results

\# in an estimate of the probability of a person being uncensored in each month.

fu_data_exp_pp %<>%

mutate(pr_cens_stab = 1 - pr_cens_stab)

\# To derive each subject’s estimated probability of their complete censoring history up to

\# each month, we multiply the estimated probabilities of being

\# uncensored for each month cumulatively over time.

fu_data_exp_pp %<>%

arrange(study_id, cummonth) %>%

group_by(study_id) %>%

mutate(pr_cens_cum_stab = cumprod(pr_cens_stab)) %>%

ungroup()

fu_data_exp_pp %<>%

mutate(pr_cens_wt_stab = pr_cens_cum_stab/pr_cens_cum)

summary(fu_data_exp_pp$pr_cens_wt_stab)

ggplot(fu_data_exp_pp, aes(x = pr_cens_wt_stab)) +

geom_histogram(binwidth = 0.005, fill = "skyblue", color = "black") +

labs(title = "Histogram of stabilised pr_cens_wt", x = "pr_cens_wt", y = "Frequency")

\## Calculate overall weight combining treatment and censoring weights

fu_data_exp_pp %<>%

mutate(rx_cens_wt = ps_w_stab * pr_cens_wt_stab)

fu_data_exp_pp %<>%

mutate(dtg_gpp = dtg)

#Fit the outcome model with IPTW and IPCW

fit_pp <- glm(cvd ~ dtg_gpp + cummonth + cummonth^2 + dtg_gpp:cummonth + dtg_gpp:cummonth^2

, data = fu_data_exp_pp,

family = quasibinomial, weights = rx_cens_wt)

#Estimate potential outcomes under treatment

fu_data_exp_pp$dtg_gpp <- 1

fu_data_exp_pp$pred1_pp <- predict(fit_pp, newdata = fu_data_exp_pp, type = "response")

#Estimate potential outcomes under control

fu_data_exp_pp$dtg_gpp <- 0

fu_data_exp_pp$pred0_pp <- predict(fit_pp, newdata = fu_data_exp_pp, type = "response")

\# make vector of max_fu month probs under treatment

pred1_max_fum_pr_pp <- fu_data_exp_pp %>%

data.frame() %>%

select(cummonth, pred1_pp) %>%

distinct() %>% # get rid of NAs

pull(pred1_pp)

\# make vector of max_fu month probs under control

pred0_max_fum_pr_pp <- fu_data_exp_pp %>%

data.frame() %>%

select(cummonth, pred0_pp) %>%

distinct() %>%

pull(pred0_pp)

#Compute standardised 3 year risk under treatment

std_rx1_var_pp <- 1- (prod(1- pred1_max_fum_pr_pp))

std_rx1_var_pp

#Compute standardised 3 year risk under control

std_rx0_var_pp <- 1- (prod(1- pred0_max_fum_pr_pp))

std_rx0_var_pp

#Compute standardised risk difference

std_risk_difference_var_pp <- std_rx1_var_pp - std_rx0_var_pp

std_risk_difference_var_pp

#Compute standardised risk ratio

std_risk_ratio_var_pp <- std_rx1_var_pp/std_rx0_var_pp

std_risk_ratio_var_pp

fu_data_exp_pp %>%

tabyl(bmi_cat)

\## BOOTSTRAP to estimate CIs in PP

\# Define a function for bootstrap

fu_data_exp_pp %<>%

mutate(dtg_gpp_boot = dtg)

output_pp <- data.frame(

boot = numeric(0),

std_rx1_var_pp_boot = numeric(0),

std_rx0_var_pp_boot = numeric(0),

std_risk_difference_var_pp_boot = numeric(0),

std_risk_ratio_var_pp_boot = numeric(0)

)

boots <- c(1:500)

#i = 1

for(i in boots){

\## Sample study_ids with replacement

in_study_ids_pp <- slice_sample(distinct(fu_data_exp_pp, study_id), prop = 1, replace = T)

\# set unique identifiers for replicated study_id's

in_study_ids_rep_pp <- in_study_ids_pp %>% # vector of study_id's as a result of sampling study_id's with replacement

group_by(study_id) %>% #filter(n()>1)

mutate(rep_study_id = paste(row_number(), study_id, sep = "_")) %>% #filter(n()>1) %>% arrange(study_id, rep_study_id) %>%

ungroup()

\# get baseline dataset for the sampled study ids

in_base_data_pp <- in_study_ids_rep_pp %>%

left_join(base_df, by = "study_id") %>%

select(-starts_with("ps"))

\# create fu data_set data for sampled study ids

in_data_pp <- in_study_ids_rep_pp %>%

left_join(fu_data_exp_pp, by = "study_id", relationship = "many-to-many") %>%

select(-starts_with("pr_cens"), -rx_cens_wt, -pred1_pp, -pred0_pp)

#Estimate the PS (use baseline data)

ps_fit_boot <- glm(dtg ~ gender + init_age + province_gp + benefit_gp + time_cat + #+ init_scheme_dur

init_cd4_cat + init_vl_cat + init_tb + init_preg + init_dm + init_htn + init_hypchol

+ init_statin , data = in_base_data_pp,

family = quasibinomial)

\# add it to the baseline dataset

in_base_data_pp$ps_bt <- fitted(ps_fit_boot)

\# calculate the INVERSE PROBABILITY of treatment weights (1/prob of treatment in treated, 1/prob of not being treated in untreated)

in_base_data_pp$ps_bt_w <- with(in_base_data_pp, dtg/ps_bt + (1-dtg)/(1-ps_bt))

\# calculate the STABILISED INVERSE PROBABILITY of treatment weights (prop exposed/prob of treatment in treated, prop unexposed/prob of not being treated in untreated)

in_base_data_pp$ps_bt_w_stab <- with(in_base_data_pp,

ifelse(dtg == 1,

mean(dtg == 1) / ps_bt,

mean(dtg == 0) / (1 - ps_bt)))

\# Add these to the follow up dataset

in_data_pp <- in_base_data_pp %>%

select(rep_study_id, ps_bt, ps_bt_w, ps_bt_w_stab) %>%

right_join(in_data_pp, by = "rep_study_id")

\### Logistic regression model to generate probability of censoring over time (including all time varying covariates)

pr_cens <- glm(cens_pp ~ init_art + gender + init_age + province_gp + benefit_gp + time_cat +

init_cd4_cat + init_vl_cat + init_tb + init_preg + init_dm + init_htn +

init_hypchol + init_statin + vl_cat + cd4_cat + preg + tb + dm + chol + statin +

cummonth, data = in_data_pp

, family = quasibinomial)

\# generate the probabilities and bind to the dataset

pr_cens_df_boot <- data.frame(predict(pr_cens, type = "response"))

pr_cens_df_boot %<>%

rename(pr_cens_boot = 1)

in_data_pp %<>%

bind_cols(pr_cens_df_boot)

in_data_pp %<>%

mutate(pr_cens_boot = 1 - pr_cens_boot) # calculate prob of being UNCENSORED

in_data_pp %<>%

arrange(rep_study_id, cummonth) %>%

group_by(rep_study_id) %>%

mutate(pr_cens_boot_cum = cumprod(pr_cens_boot)) %>%

ungroup()

\# Add unstabilised weights: 1/pr_cens_boot_cum

in_data_pp %<>%

mutate(pr_cens_boot_wt = 1/pr_cens_boot_cum)

\# add stabilised weights

\### Logistic regression model to generate probability of censoring but without time varying covariates

pr_cens_boot <- glm(cens_pp ~ init_art + gender + init_age + province_gp + benefit_gp + time_cat +

init_cd4_cat + init_vl_cat + init_tb + init_preg + init_dm + init_htn +

init_hypchol + init_statin, data = in_data_pp

, family = quasibinomial)

\# generate the probabilities and bind to the dataset

pr_cens_df_boot_stab <- data.frame(predict(pr_cens_boot, type = "response"))

pr_cens_df_boot_stab %<>%

rename(pr_cens_boot_stab = 1)

in_data_pp %<>%

bind_cols(pr_cens_df_boot_stab)

in_data_pp %<>%

mutate(pr_cens_boot_stab = 1 - pr_cens_boot_stab)

in_data_pp %<>%

arrange(rep_study_id, cummonth) %>%

group_by(rep_study_id) %>%

mutate(pr_cens_boot_cum_stab = cumprod(pr_cens_boot_stab)) %>%

ungroup()

\# calculate the stabilised weight

in_data_pp %<>%

mutate(pr_cens_boot_wt_stab = pr_cens_boot_cum_stab/pr_cens_boot_cum)

\## Calculate overall weight combining treatment and censoring weights (STABILISED)

in_data_pp %<>%

mutate(rx_cens_wt_boot = ps_bt_w_stab * pr_cens_boot_wt_stab)

#Fit the outcome model with IPTW and IPCW

model_sample <- glm(cvd ~ dtg_gpp_boot + cummonth + cummonth^2 + dtg_gpp_boot:cummonth + dtg_gpp_boot:cummonth^2#+ spline1 + spline2 + spline3 + spline4 + spline5 + spline6

, data = in_data_pp,

family = quasibinomial, weights = rx_cens_wt_boot)

#Estimate potential outcomes under treatment

in_data_pp$dtg_gpp_boot <- 1

in_data_pp$pred1_pp <- predict(model_sample, newdata = in_data_pp, type = "response")

#Estimate potential outcomes under control

in_data_pp$dtg_gpp_boot <- 0

in_data_pp$pred0_pp <- predict(model_sample, newdata = in_data_pp, type = "response")

\# make vector of max_fu month probs under treatment

pred1_max_fum_pr_pp_boot <- in_data_pp %>%

data.frame() %>%

select(cummonth, pred1_pp) %>%

distinct() %>%

pull(pred1_pp)

\# make vector of max_fu month probs under control

pred0_max_fum_pr_pp_boot <- in_data_pp %>%

data.frame() %>%

select(cummonth, pred0_pp) %>%

distinct() %>%

pull(pred0_pp)

#Compute standardised 3 year risk under treatment

std_rx1_var_pp_boot <- 1- (prod(1- pred1_max_fum_pr_pp_boot))

std_rx1_var_pp_boot

#Compute standardised 3 year risk under control

std_rx0_var_pp_boot <- 1- (prod(1- pred0_max_fum_pr_pp_boot))

std_rx0_var_pp_boot

#Compute standardised risk difference

std_risk_difference_var_pp_boot <- std_rx1_var_pp_boot - std_rx0_var_pp_boot

std_risk_difference_var_pp_boot

#Compute standardised risk ratio

std_risk_ratio_var_pp_boot <- std_rx1_var_pp_boot/std_rx0_var_pp_boot

std_risk_ratio_var_pp_boot

\# all outputs combine in one tbl

boot_output_pp <- data.frame(

boot = i,

std_rx1_var_pp_boot = std_rx1_var_pp_boot,

std_rx0_var_pp_boot = std_rx0_var_pp_boot,

std_risk_difference_var_pp_boot = std_risk_difference_var_pp_boot,

std_risk_ratio_var_pp_boot = std_risk_ratio_var_pp_boot

)

output_pp <- rbind(output_pp, boot_output_pp)

}

\# save the output to avoid regenerating

saveRDS(output_pp, file = here("output_pp.rds"))

\# Calculate the 95% confidence interval of std risk difference using percentiles

std_rx1_ci_lower_pp <- quantile(output_pp$std_rx1_var_pp_boot, 0.025)

std_rx1_ci_lower_pp

std_rx1_ci_upper_pp <- quantile(output_pp$std_rx1_var_pp_boot, 0.975)

std_rx1_ci_upper_pp

\# Calculate the 95% confidence interval of std risk ratio

std_rx0_ci_lower_pp <- quantile(output_pp$std_rx0_var_pp_boot, 0.025)

std_rx0_ci_lower_pp

std_rx0_ci_upper_pp <- quantile(output_pp$std_rx0_var_pp_boot, 0.975)

std_rx0_ci_upper_pp

\# Calculate the 95% confidence interval of std risk difference using percentiles

std_rd_ci_lower_pp <- quantile(output_pp$std_risk_difference_var_pp_boot, 0.025)

std_rd_ci_lower_pp

std_rd_ci_upper_pp <- quantile(output_pp$std_risk_difference_var_pp_boot, 0.975)

std_rd_ci_upper_pp

\# Calculate the 95% confidence interval of std risk ratio

std_rr_ci_lower_pp <- quantile(output_pp$std_risk_ratio_var_pp_boot, 0.025)

std_rr_ci_lower_pp

std_rr_ci_upper_pp <- quantile(output_pp$std_risk_ratio_var_pp_boot, 0.975)

std_rr_ci_upper_pp

\# Start capturing console output and save as output.txt

sink("output_int.txt", split = TRUE)

\# ITT

cat("ITT\n")

cat("std_rx1_var: ", std_rx1_var, "\n")

cat("std_rx1_ci_lower: ", std_rx1_ci_lower, "\n")

cat("std_rx1_ci_upper: ", std_rx1_ci_upper, "\n")

cat("std_rx0_var: ", std_rx0_var, "\n")

cat("std_rx0_ci_lower: ", std_rx0_ci_lower, "\n")

cat("std_rx0_ci_upper: ", std_rx0_ci_upper, "\n")

cat("std_risk_ratio_var: ", std_risk_ratio_var, "\n")

cat("std_rr_ci_lower: ", std_rr_ci_lower, "\n")

cat("std_rr_ci_upper: ", std_rr_ci_upper, "\n")

cat("std_risk_difference_var: ", std_risk_difference_var, "\n")

cat("std_rd_ci_lower: ", std_rd_ci_lower, "\n")

cat("std_rd_ci_upper: ", std_rd_ci_upper, "\n")

\# PP

cat("PP\n")

cat("std_rx1_var_pp: ", std_rx1_var_pp, "\n")

cat("std_rx1_ci_lower_pp: ", std_rx1_ci_lower_pp, "\n")

cat("std_rx1_ci_upper_pp: ", std_rx1_ci_upper_pp, "\n")

cat("std_rx0_var_pp: ", std_rx0_var_pp, "\n")

cat("std_rx0_ci_lower_pp: ", std_rx0_ci_lower_pp, "\n")

cat("std_rx0_ci_upper_pp: ", std_rx0_ci_upper_pp, "\n")

cat("std_risk_ratio_var_pp: ", std_risk_ratio_var_pp, "\n")

cat("std_rr_ci_lower_pp: ", std_rr_ci_lower_pp, "\n")

cat("std_rr_ci_upper_pp: ", std_rr_ci_upper_pp, "\n")

cat("std_risk_difference_var_pp: ", std_risk_difference_var_pp, "\n")

cat("std_rd_ci_lower_pp: ", std_rd_ci_lower_pp, "\n")

cat("std_rd_ci_upper_pp: ", std_rd_ci_upper_pp, "\n")

\# Stop capturing console output

sink()

### 4. R code for transition cohort sequential emulated target trial intention to treat analysis

#### 4.1 Main analysis

library(tidyverse); library(magrittr); library(lubridate); library(here)

library(base); library(janitor); library(dplyr); library(kableExtra)

library(readr); library(finalfit); library(survival); library(splines);

library(stats); library(boot); library(broom); library(data.table);

library(margins); library(tableone); library(survey)

\## ANALYSIS

\# load the dataset, one row per month per participant

tr_data <- readRDS(file = here("analysis_datasets", "tr_data.rds"))

\# Set seed for reproducibility

set.seed(123)

\## Part 1: Loop to build dataset for each trial, run analysis on that dataset, then repeat

\# make vector for number of trials

trials <- c(1:44)

\# make empty baseline dataframe with all the col headings, to which each trials baseline data will be added

all_data_base <- tr_data %>%

filter(study_id == 99999999) %>%

mutate(trial = as.numeric())

\# make empty follow up dataframe with all the col headings, to which each trials fu data will be added

all_data <- tr_data %>%

filter(study_id == 99999999) %>%

mutate(ps_w_stab = as.numeric()) %>%

mutate(cummonth_tr = as.numeric()) %>%

mutate(trial = as.numeric())

\# for i = 0 get warning message algorithm did not converge

for(i in trials){

\# i = 0

start_time <- Sys.time()

message(paste0("Trial ", i, ". Time: ", start_time))

\# make fu dataset for each trial

trial_data <- tr_data %>%

mutate(trial = i) %>%

filter(cummonth >= i & (is.na(dtg_m) | dtg_m >= i)) %>% # drop months before trial baseline (but keep baseline as need the covariate values), and drop people started on dtg before baseline

filter(trial != month_censor_itt_cvd_t) %>% #drop people who ended follow up in the baseline month (their outcomes feature in the previous trials)

group_by(study_id) %>%

mutate(cummonth_tr = cummonth - i) %>% # generate variable for follow up time within each trial

mutate(across(age:art, ~.[cummonth == i])) %>% # set all variables for each month for person to the value at baseline (as we estimate the risk of CVD per month, conditional on baseline variables)

ungroup() %>%

filter(art == 0 | art == 1) %>% # Keep only those on TEE or TLD at baseline, as 0 = "TEE" 1 = "TLD" 2= "OTHER"

filter(vl_cat != ">=1000") # drop all those with known viraemia

trial_data %<>% # get rid of excess months

filter(cummonth_tr <= 36)

\# make baseline dataset for each trial

trial_data_base <- trial_data %>%

filter(cummonth_tr == 0)

trial_data %<>%

filter(cummonth_tr != 0) # now that baseline months data is recorded in all rows for each study_id, drop the baseline month as no event can happen in it

#Estimate the PS using baseline data only

ps_fit <- glm(art ~ gender + province_gp + benefit_gp + age + haart_y_fu + #scheme_dur_y +

preg + tb + statin + vl_cat + cd4_cat + chol + dm + htn, data = trial_data_base,

family = quasibinomial)

\# add it to the dataset

trial_data_base$ps <- fitted(ps_fit)

\# calculate the STABILISED INVERSE PROBABILITY of treatment weights (prop exposed/prob of treatment in treated, prop unexposed/prob of not being treated in untreated)

trial_data_base$ps_w_stab <- with(trial_data_base,

ifelse(art == 1,

mean(art == 1) / ps,

mean(art == 0) / (1 - ps)))

\# Add these to the follow up dataset

trial_data <- trial_data_base %>%

select(study_id, ps_w_stab) %>%

right_join(trial_data, by = "study_id")

all_data <- trial_data %>%

bind_rows(all_data)

\# Bind base data to the main baseline dataframe

all_data_base <- trial_data_base %>%

bind_rows(all_data_base)

}

\# save file to avoid regenerating

saveRDS(all_data_base, file = here("analysis_datasets", "all_data_base.rds"))

saveRDS(all_data, file = here("analysis_datasets", "all_data.rds"))

\# reload to avoid regenerating

#all_data <- readRDS(file = here("analysis_datasets", "all_data.rds"))

#all_data_base <- readRDS(file = here("analysis_datasets", "all_data_base.rds"))

\# Creat table to check if weighted pseudo-population is balanced

survey_design <- svydesign(ids = ~1, data = all_data_fu_base, weights = ~ps_w_stab)

table <- svyCreateTableOne(

vars = c("age", "gender", "province_gp",

"haart_y_fu", "benefit_gp", "vl_cat",

"cd4_cat", "tb", "preg", "chol", "htn",

"dm", "statin", "bmi_cat2"), # Covariates to be included

strata = "art", # Group by treatment status

factorVars = c("tb", "preg", "chol", "htn",

"dm", "statin"),

data = survey_design	# Apply IPTW weights

)

print(table, smd = T)

########################################

\# ANALYSIS

\# make a new dtg exposure variable so that when changing the exposure as part of standardization, the original treatment allocations unaffected

all_data %<>%

mutate(art_gitt = art)

#Fit the outcome model with IPTW

fit_itt <- glm(cvd ~ art_gitt + cummonth_tr + cummonth_tr^2 + art_gitt:cummonth_tr + art_gitt:cummonth_tr^2 + trial,

data = all_data,

family = quasibinomial, weights = ps_w_stab)

#Estimate potential outcomes under treatment

all_data$art_gitt <- 1

all_data$pred1_itt <- predict(fit_itt, newdata = all_data, type = "response")

#Estimate potential outcomes under control

all_data$art_gitt <- 0

all_data$pred0_itt <- predict(fit_itt, newdata = all_data, type = "response")

\### Calculate standardised 3 year risk ratio & risk difference, allowing monthly probability to vary

\# make vector of 36 month probs under treatment

pred1_36m_pr <- all_data %>%

data.frame() %>%

select(cummonth_tr, pred1_itt, trial) %>%

distinct() %>%

filter(cummonth_tr <= 36) %>% # keep the first 36 months only

na.omit() %>%

group_by(cummonth_tr) %>%	# average the probs for each cummonth_tr

summarise(mean_pred1_itt = mean(pred1_itt)) %>%

pull(mean_pred1_itt) # make a vector of just pred1_itt

\# make vector of 36 month probs under control

pred0_36m_pr <- all_data %>%

data.frame() %>%

select(cummonth_tr, pred0_itt, trial) %>%

distinct() %>%

filter(cummonth_tr <= 36) %>% # keep the first 36 months only

na.omit() %>%

group_by(cummonth_tr) %>%	# average the probs for each cummonth_tr

summarise(mean_pred0_itt = mean(pred0_itt)) %>%

pull(mean_pred0_itt) # make a vector of just pred1_itt

#Compute standardised 3 year risk under treatment

std_rx1_var <- 1- (prod(1- pred1_36m_pr))

std_rx1_var

#Compute standardised 3 year risk under control

std_rx0_var <- 1- (prod(1- pred0_36m_pr))

std_rx0_var

#Compute standardised risk difference

std_risk_difference_var <- std_rx1_var - std_rx0_var

std_risk_difference_var

#Compute standardised risk ratio

std_risk_ratio_var <- std_rx1_var/std_rx0_var

std_risk_ratio_var

\# Start capturing console output and save as output.txt

sink("output_int_trial.txt", split = TRUE)

\# ITT

cat("ITT\n")

cat("std_rx1_var: ", std_rx1_var, "\n")

cat("std_rx0_var: ", std_rx0_var, "\n")

cat("std_risk_ratio_var: ", std_risk_ratio_var, "\n")

cat("std_risk_difference_var: ", std_risk_difference_var, "\n")

cat("std_rx1m_var: ", std_rx1m_var, "\n")

cat("std_rx0m_var: ", std_rx0m_var, "\n")

cat("std_rx1f_var: ", std_rx1f_var, "\n")

cat("std_rx0f_var: ", std_rx0f_var, "\n")

cat("std_risk_ratio_va_mr: ", std_risk_ratio_var_m, "\n")

cat("std_risk_difference_var_m: ", std_risk_difference_var_m, "\n")

cat("std_risk_ratio_var_f: ", std_risk_ratio_var_f, "\n")

cat("std_risk_difference_var_f: ", std_risk_difference_var_f, "\n")

\# Stop capturing console output

sink()

#### 4.2 Bootstrap to calculate 95% confidence intervals

library(dplyr)

library(parallel)

library(doParallel)

library(doRNG)

\## ANALYSIS

\# function that will be replicated in the bootstrap

\# it will sample a vector of study_id's from the tr_data data.frame for each iteration

tr_fnc <- function(h){

\# load the dataset

tr_data <- readRDS(file = ".../analysis_datasets/tr_data.rds")

\# sample study_id's with replacement

tr_study_ids <- slice_sample(distinct(tr_data, study_id), prop = 1, replace = T)

\# set unique identifiers for replicated study_id's and add to tr_data

tr_data <- tr_study_ids %>% # vector of study_id's as a result of sampling study_id's with replacement

group_by(study_id) %>%

mutate(rep_study_id = paste(row_number(), study_id, sep = "_")) %>%

ungroup() %>%

left_join(tr_data, by = "study_id", relationship = "many-to-many")

\## Part 1: Loop to build dataset for each trial, run analysis on that dataset, then repeat

\# make vector for number of trials

trials <- c(1:44) # Trial 0 has everyone on TLE, so models dont converge

\# make empty follow up dataframe with all the col headings, to which each trials fu data will be added

all_data <- tr_data %>%

filter(study_id == 99999999) %>%

mutate(ps_w_stab = as.numeric()) %>%

mutate(cummonth_tr = as.numeric()) %>%

mutate(trial = as.numeric())

for(i in trials){

\# make fu dataset for each trial

trial_data <- tr_data %>%

filter(cummonth >= i & (is.na(dtg_m) | dtg_m >= i)) %>% # drop months before trial baseline, and drop people started on dtg before baseline

mutate(trial = i) %>%

group_by(rep_study_id) %>%

mutate(cummonth_tr = cummonth - i) %>% # generate variable for follow up time within each trial

mutate(across(age:art, ~.[cummonth == i])) %>% # set all variables for each month for person to the value at baseline (as we estimate the risk of CVD per month, conditional on baseline variables)

ungroup() %>%

filter(art == 0 | art == 1) %>% # Keep only those on TEE or TLD at baseline, as 0 = "TEE" 1 = "TLD" 2= "OTHER")))

filter(vl_cat != ">=1000") # drop all those with known viraemia

trial_data <- trial_data %>% # get rid of excess months

filter(cummonth_tr <= 36)

\# make baseline dataset for each trial

trial_data_base <- trial_data %>%

filter(cummonth_tr == 0)

trial_data <- trial_data %>%

filter(cummonth_tr != 0) # now that baseline months data is recorded in all rows for each study_id, drop the baseline month as no event can happen in it

#Estimate the PS using baseline data only

ps_fit <- glm(art ~ gender + province_gp + benefit_gp + age + haart_y_fu + # scheme_dur_y +

preg + tb + statin + vl_cat + cd4_cat + chol + dm + htn, data = trial_data_base,

family = quasibinomial)

\# add it to the dataset

trial_data_base$ps <- fitted(ps_fit)

\# calculate the STABILISED INVERSE PROBABILITY of treatment weights (prop exposed/prob of treatment in treated, prop unexposed/prob of not being treated in untreated)

trial_data_base$ps_w_stab <- with(trial_data_base,

ifelse(art == 1,

mean(art == 1) / ps,

mean(art == 0) / (1 - ps)))

\# Add these to the follow up dataset

trial_data <- trial_data_base %>%

select(rep_study_id, ps_w_stab) %>%

right_join(trial_data, by = "rep_study_id")

all_data <- trial_data %>%

bind_rows(all_data)

} # for loop

\# get rid of unnecessary variables

all_data <- all_data %>%

select(rep_study_id, gender, cvd, art, cummonth, cummonth_tr, ps_w_stab, trial)

\# remove unused datasets and clear memory.

#rm(all_data)

rm(tr_data)

rm(trial_data)

rm(trial_data_base)

rm(ps_fit)

rm(tr_study_ids)

\# make a new dtg exposure variable so that when changing the exposure as part of standardization, the original treatment allocations unaffected

all_data <- all_data %>%

mutate(art_gitt = art)

#Fit the outcome model with IPTW

fit_itt <- glm(cvd ~ art_gitt + cummonth_tr + cummonth_tr^2 + art_gitt:cummonth_tr + art_gitt:cummonth_tr^2 + trial,# + spline1 + spline2 + spline3 + spline4 + spline5 + spline6,

data = all_data,

family = quasibinomial, weights = ps_w_stab)

#Estimate potential outcomes under treatment

all_data$art_gitt <- 1

all_data$pred1_itt <- predict(fit_itt, newdata = all_data, type = "response")

#Estimate potential outcomes under control

all_data$art_gitt <- 0

all_data$pred0_itt <- predict(fit_itt, newdata = all_data, type = "response")

\### Calculate standardised 3 year risk ratio & risk difference, allowing monthly probability to vary

\# make vector of 36 month probs under treatment

pred1_36m_pr <- all_data %>%

data.frame() %>%

select(cummonth_tr, pred1_itt, trial) %>%

distinct(cummonth_tr, pred1_itt, .keep_all = T) %>%

filter(cummonth_tr <= 36) %>%

na.omit() %>%

group_by(cummonth_tr) %>%

summarise(mean_pred1_itt = mean(pred1_itt)) %>%

pull(mean_pred1_itt)

\# make vector of 36 month probs under control

pred0_36m_pr <- all_data %>%

data.frame() %>%

select(cummonth_tr, pred0_itt, trial) %>%

distinct(cummonth_tr, pred0_itt, .keep_all = T) %>%

filter(cummonth_tr <= 36) %>%

na.omit() %>%

group_by(cummonth_tr) %>%

summarise(mean_pred0_itt = mean(pred0_itt)) %>%

pull(mean_pred0_itt)

#Compute standardised 3 year risk under treatment

std_rx1_var <- 1- (prod(1- pred1_36m_pr))

std_rx1_var

#Compute standardised 3 year risk under control

std_rx0_var <- 1- (prod(1- pred0_36m_pr))

std_rx0_var

#Compute standardised risk difference

std_risk_difference_var <- std_rx1_var - std_rx0_var

std_risk_difference_var

#Compute standardised risk ratio

std_risk_ratio_var <- std_rx1_var/std_rx0_var

std_risk_ratio_var

\# outcome of trial combine in one tbl

tibble(

boot = h,

std_rx1_var = std_rx1_var,

std_rx0_var = std_rx0_var,

std_risk_difference_var = std_risk_difference_var,

std_risk_ratio_var = std_risk_ratio_var,

)

} # end of function

\# set the number of clusters appropriate for the machine it is running on

my_cluster <- makeCluster(detectCores() - 4)

clusterEvalQ(my_cluster, library(dplyr))

\# for reproducability

registerDoParallel(my_cluster)

registerDoRNG(123)

\# run the bootstrap 500 times on all_data

start <- Sys.time()

try(outcome_par_lapply <- clusterApply(my_cluster, # the clusters

c(1:500), # the list to iterate over

tr_fnc)

)

stopCluster(my_cluster)

end <- Sys.time()

\# # Calculate the amount of time to run all the trials

end - start

\# put the results of all the trials in a readable shape

tbl_outcome_par <- bind_rows(outcome_par_lapply)

tbl_outcome_par

saveRDS(tbl_outcome_par, ".../tbl_outcome_par_lapply.rds")

\# reload to save calculating each time

#tbl_outcome_par <- readRDS(file = ".../tbl_outcome_par_lapply.rds")

tibble(bootstraps = nrow(tbl_outcome_par),

start = start,

end = end) %>%

mutate(total = end-start) %>%

saveRDS(".../boot_strap_times_parlapply.rds")

\# Calculate the 95% confidence interval of std risk under treatment using percentiles

std_rx1_ci_lower <- quantile(tbl_outcome_par$std_rx1_var, 0.025)

std_rx1_ci_lower

std_rx1_ci_upper <- quantile(tbl_outcome_par$std_rx1_var, 0.975)

std_rx1_ci_upper

\# Calculate the 95% confidence interval of std risk under control

std_rx0_ci_lower <- quantile(tbl_outcome_par$std_rx0_var, 0.025)

std_rx0_ci_lower

std_rx0_ci_upper <- quantile(tbl_outcome_par$std_rx0_var, 0.975)

std_rx0_ci_upper

\# Calculate the 95% confidence interval of std risk difference using percentiles

std_rd_ci_lower <- quantile(tbl_outcome_par$std_risk_difference_var, 0.025)

std_rd_ci_lower

std_rd_ci_upper <- quantile(tbl_outcome_par$std_risk_difference_var, 0.975)

std_rd_ci_upper

\# Calculate the 95% confidence interval of std risk ratio using percentiles

std_rr_ci_lower <- quantile(tbl_outcome_par$std_risk_ratio_var, 0.025)

std_rr_ci_lower

std_rr_ci_upper <- quantile(tbl_outcome_par$std_risk_ratio_var, 0.975)

std_rr_ci_upper

cis <- tibble(

std_rx1_ci_lower = std_rx1_ci_lower,

std_rx1_ci_upper = std_rx1_ci_upper,

std_rx0_ci_lower = std_rx0_ci_lower,

std_rx0_ci_upper = std_rx0_ci_upper,

std_rd_ci_lower = std_rd_ci_lower,

std_rd_ci_upper = std_rd_ci_upper,

std_rr_ci_lower = std_rr_ci_lower,

std_rr_ci_upper = std_rr_ci_upper

)

saveRDS(cis, ".../cis_nospline.rds")
